# Theta-Beta Ratio in Attention Deficit Hyperactivity Disorder: A Multiverse Analysis

**DOI:** 10.64898/2026.01.08.26343676

**Authors:** Dawid Strzelczyk, Andrea Vetsch, Nicolas Langer

## Abstract

Attention Deficit Hyperactivity Disorder (ADHD) affects 5-7% of children worldwide, yet diagnosis continues to rely on clinical-behavioral assessments. The theta/beta ratio (TBR) derived from electroencephalography (EEG) has long been proposed as a complementary neurobiological marker of ADHD, based on reports of elevated TBR in affected children. However, accumulating evidence has raised concerns about the robustness and generalizability of these findings, pointing to a strong sensitivity to methodological choices. Here, we used multiverse analyses to systematically quantify how researcher degrees of freedom shape conclusions about associations between TBR and ADHD. Across two large, independent datasets (Healthy Brain Network: N=1,499; validation sample: N=381), we evaluated 576 of theoretically plausible analytical specifications, varying recording conditions, reference scheme, frequency band definitions, treatment of aperiodic (1/f) activity, regions of interest, sample inclusion criteria and covariate specifications. Across the multiverse, we found that group differences in TBR were highly contingent on analytical choices, with no evidence for robust main effects of diagnosis, indicating no reliable differences between healthy controls, ADHD-inattentive, and ADHD-combined subtypes. Instead, significant effects emerged primarily as interactions with age and individual alpha frequency (IAF), particularly when TBR was derived from aperiodic-uncorrected power or from the aperiodic signal itself. These interaction patterns replicated across both independent samples and were observed using both categorical and dimensional definitions of ADHD. Together, these findings indicate that previously reported TBR effects are largely driven by variability in aperiodic activity and IAF rather than genuine differences in oscillatory theta-beta dynamics. Our results challenge the interpretation of TBR as a reliable standalone biomarker for ADHD and underscore the importance of multiverse approaches for evaluating candidate neurobiological markers in heterogeneous clinical populations.

## 1. Introduction

Attention Deficit Hyperactivity Disorder (ADHD) affects approximately 5-7% of children worldwide and represents one of the most prevalent neurodevelopmental disorders, with significant implications for public health and educational systems (Boxum et al., 2024; Polanczyk et al., 2007; Sinishaw et al., 2024). Core symptoms such as inattention, hyperactivity, and impulsivity can significantly impair academic performance, disrupt social relationships, and increase the risk of comorbid mental health conditions, with these challenges frequently persisting into adulthood (Arns et al., 2013; Boxum et al., 2024; Faraone et al., 2006). Current diagnostic practices rely heavily on subjective clinical assessments, behavioral rating scales, and structured interviews, creating a pressing need for objective neurobiological markers that could enhance diagnostic precision and reduce the substantial variability in clinical decision-making (Young et al., 2008).

Numerous studies have shown that children with ADHD often exhibit increased slow-wave activity, particularly in the theta range (4-8 Hz), alongside decreased beta activity (>14 Hz) (Arns et al., 2013; Bresnahan et al., 1999; Callaway et al., 1983; Lubar, 1991; Mann et al., 1992; Matsuura et al., 1993). In the context of ADHD, increased theta has been associated with fatigue and drowsiness, whereas lower beta has been linked to decreased mental activity and concentration (Boxum et al., 2024; Loo & Arns, 2015). To quantify this imbalance, Lubar (1991) proposed the theta/beta ratio (TBR) as a potential biomarker of ADHD, suggesting it could serve as a reliable tool for both diagnosis and treatment monitoring. This approach gained traction with evidence from Monastra et al. (1999), who reported high sensitivity (86%) and specificity (98%) for classifying ADHD in a multi-center study (Monastra et al., 1999, 2001). These promising results contributed to regulatory recognition, culminating in the U.S. FDA’s approval of TBR-based assessment as an adjunctive tool for ADHD diagnosis (Arns et al., 2016; Dolgin, 2014; Snyder et al., 2008, 2015). Consequently, multiple commercial EEG-based diagnostic systems emerged, promising to enhance the objectivity and reliability of ADHD evaluations in clinical practice.

However, despite early promising results, more recent studies with heterogeneous samples have raised concerns about the validity and robustness of TBR as a diagnostic marker for ADHD (Arns et al., 2013, 2016; Boxum et al., 2024; Lenartowicz & Loo, 2014; Loo & Arns, 2015; Saad et al., 2018; van Dijk et al., 2020). As pointed out by Arns et al. (2013, 2016), the foundational studies by Monastra and colleagues (1999, 2001) contained several methodological limitations that may compromise their generalizability. Both studies employed unbalanced designs with ADHD groups approximately 5 times larger than controls, potentially inflating statistical power (Brysbaert, 2019; Ioannidis, 2005). While manual artifact rejection was performed, no specific thresholds or rejection rates were reported across groups, a critical omission given that movement artifacts significantly affect the aperiodic (1/f) component of EEG and can substantially alter TBR values (Donoghue, 2025; Donoghue, Dominguez, et al., 2020; Donoghue, Haller, et al., 2020; Karalunas et al., 2022; Schmidt et al., 2025; Tröndle & Langer, 2026b). The risk of biased TBR estimates is particularly high, because children with ADHD tend to move more than controls (Dziemian et al., 2022). Furthermore, the ADHD samples excluded comorbid conditions such as anxiety, depression, or learning disorders, despite these being present in 60-80% of real-world ADHD cases, severely limiting ecological validity (Ioannidis, 2005; Polanczyk et al., 2007; Sinishaw et al., 2024). Importantly, the authors conflated statistically significant group differences with diagnostic utility, yet demonstrating higher average TBR in ADHD groups does not establish the measure’s capacity for reliable individual classification.

Meta-analyses by Arns et al. (2013, 2016) have revealed significant heterogeneity across studies and a marked decline in effect sizes over time, indicating that more recent studies tend to find smaller or non-significant differences in TBR between individuals with ADHD and healthy controls. This growing inconsistency has led to the view that TBR may only be a reliable marker of ADHD under specific methodological conditions. Factors such as preprocessing pipelines, resting state conditions (eyes open vs. eyes closed), reference schemes, power computation methods, frequency band definitions, regions of interest, and participant characteristics (e.g., medication status, comorbidities) can all influence the resulting TBR values (Kerson et al., 2020; van Dijk et al., 2020). Supporting this, van Dijk et al. (2020) processed the same dataset with five alternative pipelines that varied aperiodic (1/f) handling, spectral estimation (i.e., Welch, Multi-taper, Wavelet), and the TBR formula. Although absolute TBR values differed significantly, the estimates were highly correlated and none reliably distinguished ADHD from controls. However, five pipelines cover only a small fraction of plausible specifications, and other reasonable choices could similarly shift the estimates.

The multitude of choices researchers face during data preprocessing and analysis is referred to as researcher degrees of freedom (RDF) (Simmons et al., 2011). When these decisions remain undisclosed or are made post-hoc based on desired outcomes, they can inflate false-positive rates and undermine the reproducibility of scientific findings (Götz et al., 2024; Simmons et al., 2011). To address these concerns, the scientific community has increasingly embraced multiverse analysis as a robust framework for evaluating the stability of research findings across the full spectrum of reasonable analytical choices (Del Giudice & Gangestad, 2021; Götz et al., 2024; Sarma et al., 2024, 2025; Steegen et al., 2016). Rather than presenting results from a single, potentially arbitrary analytical pathway, multiverse analysis systematically implements all plausible combinations of methodological decisions, providing a comprehensive assessment of effect robustness and identifying specific conditions under which effects emerge or disappear (Del Giudice & Gangestad, 2021; Sarma et al., 2025; Steegen et al., 2016).

Therefore, in the present study, we employed a multiverse analysis to systematically address the methodological uncertainties surrounding the computation of TBR in children and adolescents with ADHD and healthy controls. Using two large, independent samples: the Healthy Brain Network (N = 1499) and a validation sample (N = 381), we systematically varied preprocessing steps and TBR computation methods to evaluate the consistency of observed group differences across the full reasonable analytical space. Our choice of analytic specifications was informed by a systematic review of prior work on TBR in ADHD (see Methods 2.6). This comprehensive framework enabled us to determine whether TBR differences between children and adolescents with ADHD and healthy controls represent robust neurobiological phenomena or are effects of specific methodological choices, thereby providing crucial evidence for the field’s ongoing evaluation of EEG-based ADHD biomarkers.

## 2. Methods

To ensure transparency and reproducibility, all analysis code, EEG features, and demographic data used in the statistical models are accessible on OSF via https://osf.io/u5yxv.

### 2.1. Participants

To assess the generalizability of our findings, we analyzed data from two independent samples: a main sample from the Healthy Brain Network (HBN) project (Alexander et al., 2017; Langer et al., 2017) and a validation sample taken from a large-scale multicenter clinical study (Müller et al., 2020; Münger et al., 2021, 2022). As the multiverse analysis results were highly similar, and preprocessing and data analysis were identical across samples, we report demographic details, EEG acquisition information and all results for the validation sample in the Supplement (Supplementary Material 1.2).

The HBN project by the Child Mind Institute is an ongoing initiative that aims to generate a freely available biobank of multimodal datasets of children, adolescents and young adults aged 5-22 years. For the current study, we analyzed data available up to Release 11 (as of 11.23.2022), identifying a total of 1,499 participants with resting state EEG and either no psychiatric diagnosis (healthy controls) or a diagnosis of ADHD. The initial sample before all exclusions consisted of 271 healthy controls (mean age = 9.81 years, SD = 3.52, range = 5.02-21.67), 584 participants with ADHD-Combined Type (mean age = 9.19 years, SD = 2.86, range = 5.04-21.72), and 559 with ADHD-Inattentive Type (mean age = 10.88 years, SD = 3.07, range = 5.43-21.48). An additional 85 participants with ADHD-Hyperactive/Impulsive Type (mean age = 7.65 years, SD = 2.10, range = 5.01-13.67) were excluded from further analyses due to insufficient sample size across key subgroup factors used in the multiverse analysis (e.g., gender, comorbidity and medication status, and their interactions), which were central to the multiverse analysis framework. For detailed demographic information please refer to Table 1.

**Table 1.**
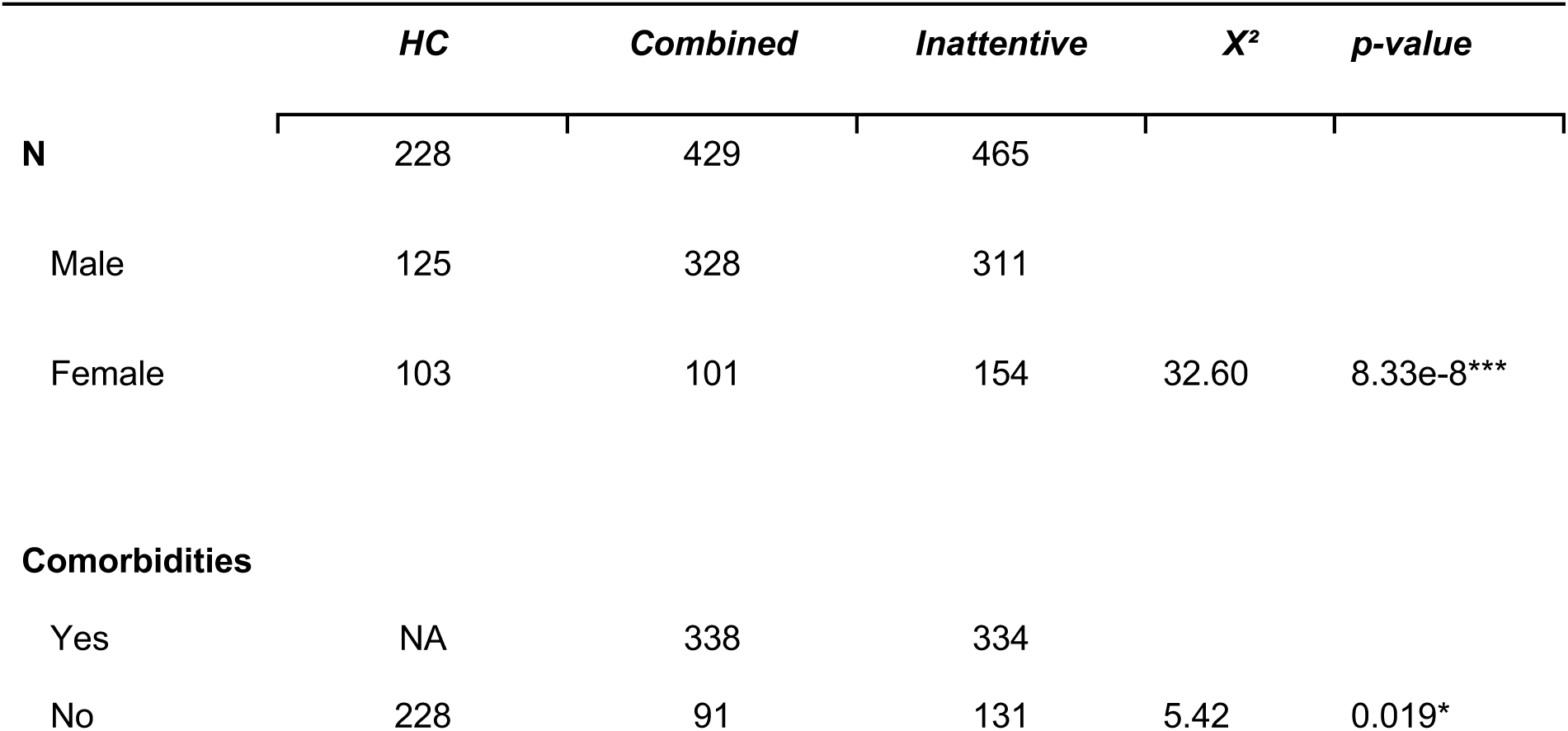
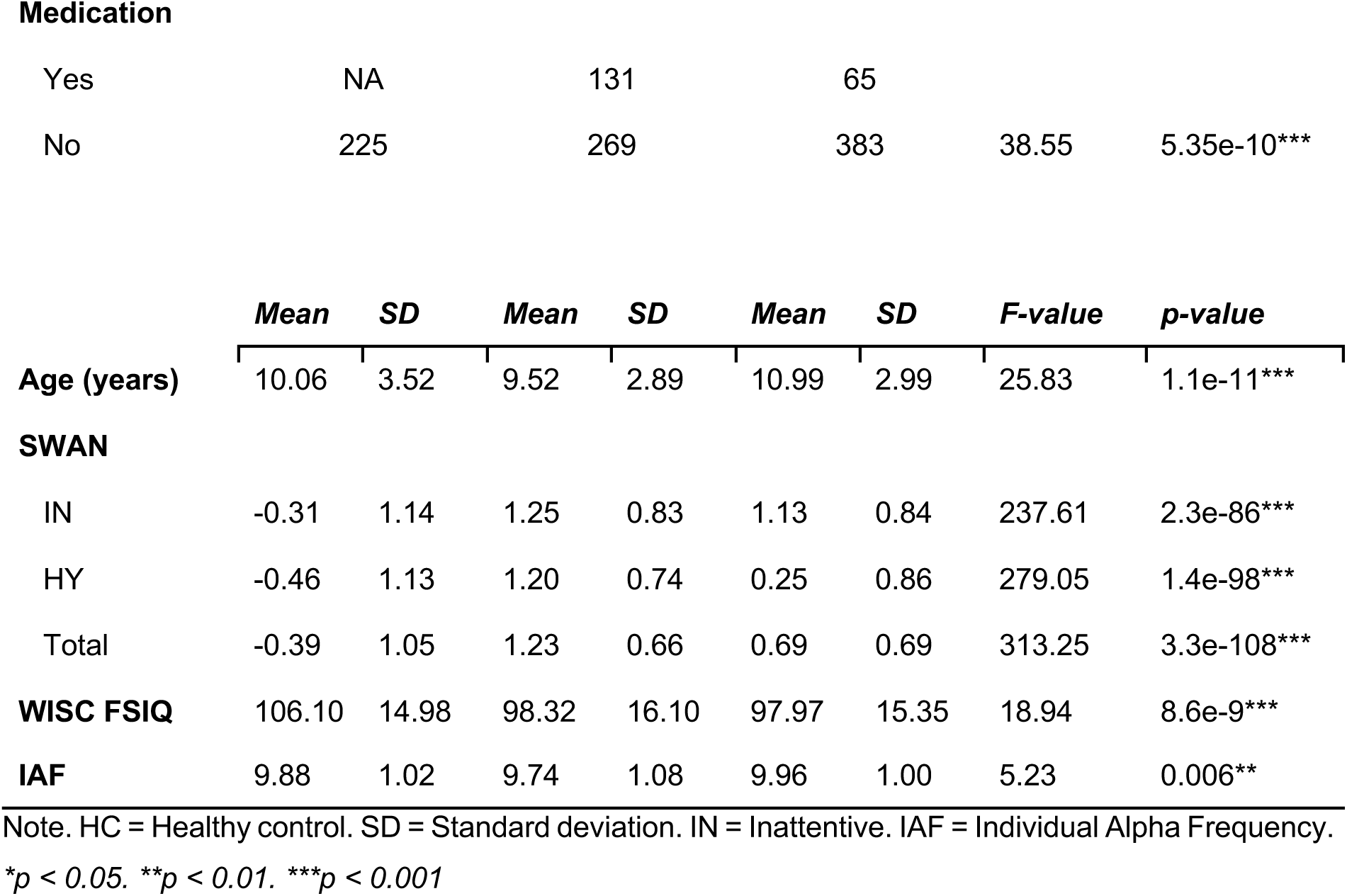
Demographics information of the Healthy Brain Network sample.

|  | *HC* | *Combined* | *Inattentive* | *X<sup>2</sup>* | *p-value* |
| --- | --- | --- | --- | --- | --- |
| **N** | 228 | 429 | 465 |  |  |
| Male | 125 | 328 | 311 |  |  |
| Female | 103 | 101 | 154 | 32.60 | 8.33e-8*** |
| **Comorbidities** |  |  |  |  |  |
| Yes | NA | 338 | 334 |  |  |
| No | 228 | 91 | 131 | 5.42 | 0.019* |
| **Medication** |  |  |  |  |  |
| Yes | NA | 131 | 65 |  |  |
| No | 225 | 269 | 383 | 38.55 | 5.35e-10*** |

|  | *Mean* | *SD* | *Mean* | *SD* | *Mean* | *SD* | *F-value* | *p-value* |
| --- | --- | --- | --- | --- | --- | --- | --- | --- |
| **Age (years)** | 10.06 | 3.52 | 9.52 | 2.89 | 10.99 | 2.99 | 25.83 | 1.1e-11*** |
| **SWAN** |  |  |  |  |  |  |  |  |
| IN | -0.31 | 1.14 | 1.25 | 0.83 | 1.13 | 0.84 | 237.61 | 2.3e-86*** |
| HY | -0.46 | 1.13 | 1.20 | 0.74 | 0.25 | 0.86 | 279.05 | 1.4e-98*** |
| Total | -0.39 | 1.05 | 1.23 | 0.66 | 0.69 | 0.69 | 313.25 | 3.3e-108*** |
| **WISC FSIQ** | 106.10 | 14.98 | 98.32 | 16.10 | 97.97 | 15.35 | 18.94 | 8.6e-9*** |
| **IAF** | 9.88 | 1.02 | 9.74 | 1.08 | 9.96 | 1.00 | 5.23 | 0.006** |

Note. HC = Healthy control. SD = Standard deviation. IN = Inattentive. IAF = Individual Alpha Frequency.
\* $p < 0.05$ . \*\* $p < 0.01$ . \*\*\* $p < 0.001$

ADHD diagnoses in the HBN study were determined by licensed clinicians based on a structured clinical protocol (Alexander et al., 2017). The primary diagnostic instrument was the Kiddie Schedule for Affective Disorders and Schizophrenia (K-SADS; (Chambers et al., 1985)), a semi-structured interview conducted with parents and, for participants aged 11 and older, also with the child. Additional clinical judgment was informed by behavioral observations during testing sessions, family history of psychiatric conditions, prior diagnoses, and responses on standardized parent-, child-, and teacher-report questionnaires (e.g., CBCL (Achenbach & Dumenci, 2001), SWAN (Swanson et al., 2001), Conners (Conners, 2008)). The decision also took into account whether the participant had previously received mental health support. For detailed information, please refer to the HBN Diagnostic Process (Version 3): (https://fcon_1000.projects.nitrc.org/indi/cmi_healthy_brain_network). The IQ was measured using the Wechsler Intelligence Scale for Children (WISC-V), Wechsler Abbreviated Scale of Intelligence (WASI), or Wechsler Adult Intelligence Scale (WAIS) according to the age of the participant. 280 ADHD participants were regularly treated with psychiatric medication in daily life (i.e., ADHD stimulants (Methylphenidate-based: Ritalin, Concerta; Amphetamine-based: Adderall, Vyvanse), Antidepressants (Prozac, Zoloft) and atypical antipsychotics (Abilify)). Prior to participation, legal guardians or participants of legal age provided written informed consent. Study approval was given by the Chesapeake Institutional Review Board.

### 2.2. EEG Acquisition

The HBN EEG data were recorded using a 128-channel Hydrogel net system (Electrical Geodesics Inc.) at a sampling rate of either 250 Hz or 500 Hz. To ensure consistency across recordings, all EEG data were downsampled to 250 Hz prior to further processing. The reference electrode was placed at Cz (vertex of the head), and electrode impedances were kept below 40 kΩ, lower than EGI’s standard recommendation of 50 (Net Station Acquisition Technical Manual).

Participants from the HBN sample were seated comfortably in a sound-shielded room, 70 cm away from a 17-inch CRT monitor (Sony Trinitron Multiscan G220; display: 330 × 240 mm; resolution: 800 × 600 pixels; refresh rate: 100 Hz). A chinrest was used to minimize head movements. Participants were informed that EEG would be recorded while they alternated between eyes-open (EO) and eyes-closed (EC) resting-state conditions. Task instructions were presented on the screen, and a research assistant provided clarification via intercom from an adjacent control room. Compliance was monitored through a live video feed. The resting state task consisted of five EO-EC cycles. Each EO block lasted 20 s (totaling 1 min 40 s), and each EC block lasted 40 s (totaling 3 min 20 s), resulting in a total recording time of 5 minutes. Pre-recorded verbal cues were delivered via loudspeakers to instruct participants when to open or close their eyes. During EO blocks, participants were asked to maintain gaze on a central fixation cross. The EO-EC alternation aimed to prevent fatigue and maintain vigilance, with longer EC periods to capitalize on lower artifact rates in the EC condition.

### 2.3. EEG Preprocessing

EEG data from both the main and validation datasets were preprocessed using an identical pipeline to ensure methodological consistency across samples. Data preprocessing and EEG feature extraction were conducted using MATLAB 2023b (The MathWorks, Inc., Natick, MA, USA) and RStudio 4.4.1 (R Core Team). The data were preprocessed in Automagic 3.1, a MATLAB based toolbox for automated, reliable and objective preprocessing of EEG datasets (Pedroni et al., 2019). In a first step, the bad channels were detected using the PREP pipeline (Bigdely-Shamlo et al., 2015). A channel was defined as bad based on 1) extreme amplitudes (z-score cutoff for robust channel deviation of more than 5), 2) lack of correlation (at least 0.4) with other channels with a window size of 1s to compute the correlation, 3) lack of predictability by other channels (channel is bad if the prediction falls below the absolute correlation of 0.75 in a fraction of 0.4 windows of a duration of 5s), 4) unusual high frequency noise using a z-score cutoff for SNR of 5. These channels were removed from the original EEG data. The data was filtered using a high-pass filter with 0.5 Hz cutoff using the EEGLAB function pop_eegfiltnew (Widmann & Schröger, 2012). Line noise was removed using a ZapLine method with a passband edge of 60 Hz for HBN sample and 50 Hz for validation sample (de Cheveigné, 2020), removing 7 power line components. Next, independent component analysis (ICA) was performed. However, as the ICA is biased towards high amplitude and low frequency noise (i.e., sweating), the data was temporarily filtered with a high-pass filter of 2 Hz in order to improve the ICA decomposition. Using the pre-trained classifier IClabel (Pion-Tonachini et al., 2019) each independent component with a probability rating >0.8 of being an artifact such as muscle activity, heart artifacts, eye activity, line noise and channel noise were removed from the data. The remaining components were back-projected on the original 0.5 Hz high-pass filtered data. In the next step, the channels identified as bad were interpolated using the spherical interpolation method. Finally, the quality of the data was automatically and objectively assessed in Automagic, thus increasing research reproducibility by having objective measures for data quality. Using a selection of 4 quality measures, the data was classified into three categories: “good”, “ok” or “bad”. Data was classified as “bad”, if 1) the proportion of high-amplitude data points (>30μV) in the signal is greater than 0.3, or 2) more than 50% of time points show a variance greater than 50μV across all channels, or 3) 30% of the channels show variance greater than 40μV, or 4) the ratio of bad channels is greater than 0.3. For the further analysis only the datasets with “good” and “ok” ratings were used. In the HBN sample, classification was performed at the subject level (1,020 “good”, 430 “ok”, 45 “bad”; 45 subjects excluded) because EO and EC blocks were recorded as alternating segments within a single continuous file per participant. In the validation dataset, classification was performed at the file level (521 “good”, 230 “ok”, 12 “bad”; 12 files excluded) because EO and EC were recorded and stored as separate files.

Subsequently, in the HBN sample, which was recorded using a 128-channel Hydrogel net system, 23 channels were excluded from further analysis: 10 EOG channels and 13 channels located on the chin and neck, as they capture minimal brain activity and are typically contaminated with muscle artifacts (Langer et al., 2017; Tröndle et al., 2020, 2021).

Next, the data from both samples was re-referenced either to average reference or linked mastoid (see multiverse analysis tree) into 2 s long segments. The first and the last segment of each EO / EC block was discarded to exclude motor activity related to opening and closing the eyes and auditory activity due to the prompt from the speakers. Remaining EEG segments were inspected using an amplitude threshold of 90 μV. In the HBN sample, 197 subjects (12.6%) were excluded from further analysis because more than 60% of their trials exceeded this threshold. Among the remaining subjects, an average of 18% of trials were excluded based on this criterion. In the validation sample, 2 subjects (0.5%) were excluded for the same reason, with an average of 14% of trials removed among the remaining subjects.

### 2.5. Feature Extraction

The features were extracted from EO and EC conditions, using both average and linked mastoid references. For each condition and reference combination, we computed three types of features: relative power, aperiodic-adjusted power, and the aperiodic component. Spectral analysis was performed using the Fast Fourier Transform (FFT) over the 1-40 Hz range after applying a single Hanning window, implemented in Fieldtrip (Oostenveld et al., 2011). First, to obtain relative power, total power spectra were computed using cfg.output = ‘pow’. These power spectra were then normalized by dividing each frequency bin by the mean power across all bins, yielding relative power estimates. Next, to compute aperiodic-adjusted power, the power spectra were decomposed into periodic and aperiodic components using the SpecParam algorithm (Donoghue, Haller, et al., 2020), implemented in FieldTrip (i.e., cfg.output = ‘fooof’). SpecParam was applied over the 1-40 Hz frequency range to minimize the risk of overfitting low-frequency noise as narrowband peaks. The algorithm settings were as follows: peak width limits set to [1, 8], maximum number of peaks of 6, minimum peak height of 0, border threshold of 5, peak threshold of 2 standard deviations above the mean, and the aperiodic mode set to ‘fixed’. The aperiodic component was reconstructed based on its fitted parameters and subtracted from the total power spectrum in log10 power space, resulting in an aperiodic-adjusted, 1/f-corrected power spectrum. The resulting values were then transformed back to linear scale and therefore represent power relative to the estimated aperiodic background. To ensure data quality, we applied a model fit threshold of R² = 0.85, calculated as the mean R² across all subjects minus 2.5 SD, following established procedures from the seminal specParm publication (Donoghue, Haller, et al., 2020; Finley et al., 2022). A total of 69 subjects from the HBN sample were excluded from further analyses due to poor model fit. For the remaining participants, the mean goodness-of-fit (R²) of the aperiodic decomposition was 0.97 ± 0.02 (M ± SD). In the validation sample, 40 subjects were excluded due to poor model fit, with remaining subjects showing R² of 0.97 ± 0.03 (M ± SD). Summarized, we obtained 6 datasets per participant: EC and EO data, each with relative power (i. e., uncorrected power), aperiodic-adjusted power (i. e., 1/f corrected), and the isolated aperiodic component (i. e., aperiodic signal only).

Subsequently, we proceeded with the computation of the individual alpha peak frequency (IAF) as the shift of the IAF during brain maturation might introduce a bias when power is averaged within a fixed-frequency window (Corcoran et al., 2018; Tröndle et al., 2022a). Specifically, it has been shown that the IAF increases over children’s development, and the rate of increase can vary among individuals (Tröndle et al., 2022a). To account for this, we estimated IAF from aperiodic-adjusted power spectra obtained during EC recordings over posterior electrodes (HBN: E62 (Pz), E75 (Oz), E70 (O1), E83 (O2), E72, E71, E76; Validation Sample: P7/T5, P3, O1, Pz, O2, P4, P8/T6). IAF was defined as the frequency of maximum power within a pre-specified range of 7 to 14 Hz, consistent with prior work (Babiloni et al., 2020; Klimesch, 1999; Tröndle et al., 2022b). If the peak was located outside of the preselected frequency range or at the border, no alpha peak was extracted for that subject, and the corresponding data was excluded from all further analyses. In the HBN sample, IAF could not be identified in 13 subjects. For the remaining participants, the mean IAF (M ± SD) was 9.74 ± 1.08 Hz in the ADHD-Combined group, 9.96 ± 1.00 Hz in the ADHD-Inattentive group, and 9.88 ± 1.02 Hz in the group without a diagnosis. In the validation sample, IAF could not be determined in 10 subjects. For the remaining participants, the mean IAF was 9.58 ± 1.08 Hz in the ADHD-Combined group, 9.60 ± 1.11 Hz in the ADHD-Inattentive group and 9.82 ± 1.06 Hz in the healthy control group.

Subsequently, theta and beta power were computed for each participant, condition, and spectral estimate using two approaches: canonical frequency bands and individualized frequency bands. Canonical bands were defined as 4-8 Hz for theta and 13-30 Hz for beta (Klimesch, 2012). Individualized bands were centered on the IAF, defined as theta = IAF-6 Hz to IAF-4 Hz, and beta = IAF+2 Hz to 30 Hz. The selection of these frequency bands is grounded in the seminal work of Wolfgang Klimesch (1999), who demonstrated that the alpha band can be divided into distinct lower and upper sub-bands. The lower alpha band extends up to 4 Hz below the IAF, covering a broader range of approximately 3.5 to 4 Hz, while the upper alpha band, which lies above the IAF, is narrower, spanning about 1 to 1.5 Hz. Klimesch also characterized the theta band as a frequency range that is approximately 2 Hz below the lower alpha band (Klimesch, 1999, 2012). The 2020 guidelines from the International Federation of Clinical Neurophysiology (IFCN) reaffirmed Klimesch’s division of the alpha and theta bands (Babiloni et al., 2020). Finally, TBR was calculated by dividing theta power by beta power (Lubar, 1991). TBR values were extracted from multiple scalp regions of interest (ROIs), selected based on prior literature (Arns et al., 2013, 2016, 2018; Boxum et al., 2024; Lenartowicz & Loo, 2014; Loo et al., 2013; Loo & Arns, 2015; Lubar, 1991; Monastra et al., 1999). These ROIs are illustrated in Figure 1.

**Figure 1.**
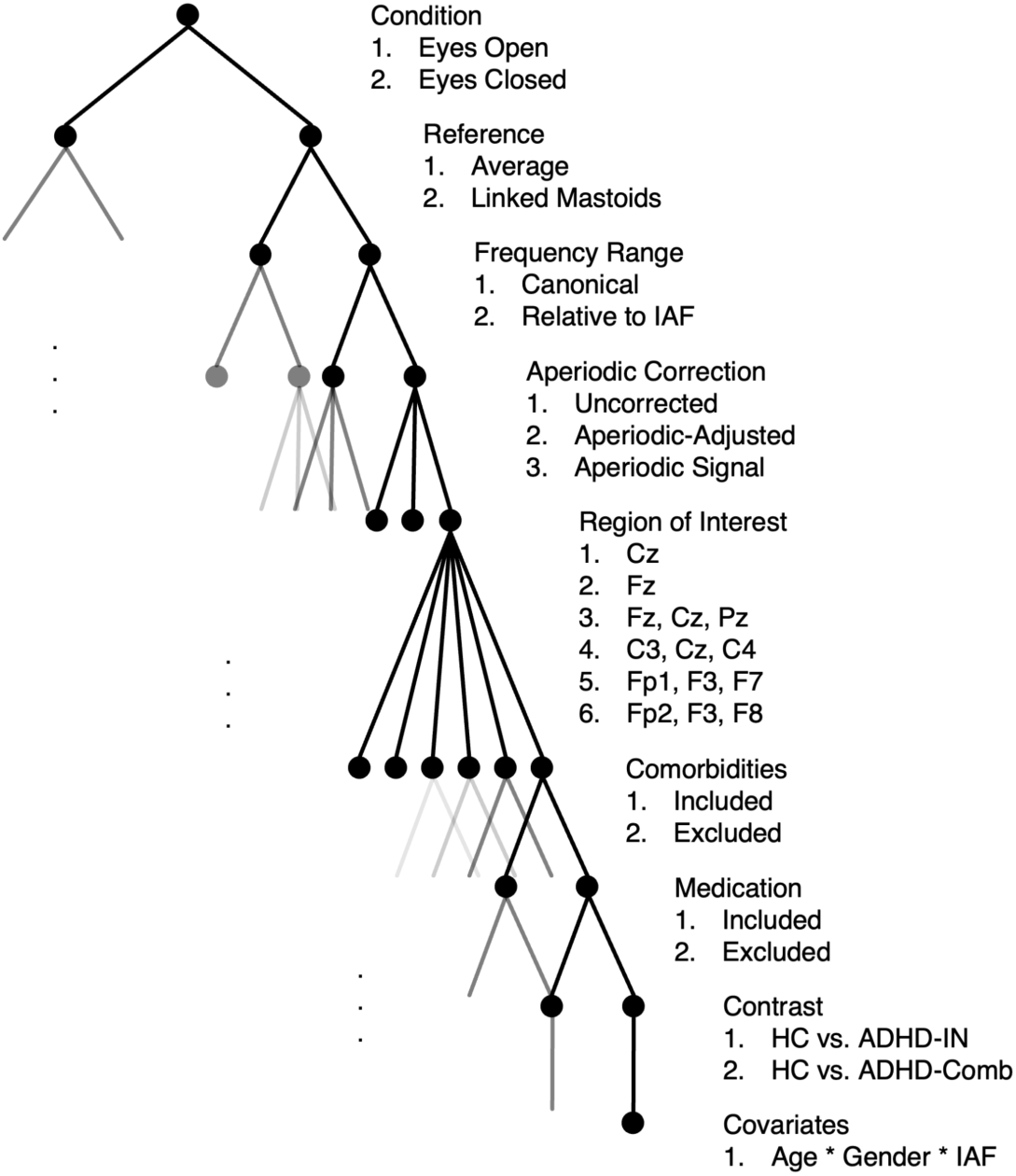
Final analysis tree illustrating all included analytical specifications. Each node represents a distinct analytical parameter in the multiverse analysis. Note that due to space limitations, not all parameter nodes are displayed in this visualization. Information on comorbid diagnoses and medication status was available only in the HBN sample, and these factors were therefore investigated exclusively in this dataset. HC = Healthy Control, IAF = Individual Alpha Frequency.

### 2.6. Multiverse Analyses

Multiverse analysis was conducted in RStudio (R version 4.4.1) using the multiverse package (version 0.6.1; Sarma et al., 2021). The multiverse framework differs from simply repeating the same statistical analysis multiple times, because it first requires the researcher to define a structured analysis space consisting of multiple defensible analytic decisions. These decisions are then expanded into all valid combinations, with each combination representing one complete analysis specification, or “universe”, providing a transparent and reproducible record of which analytic decisions were considered and how they were combined. In addition, the package reduces the need to manually write, modify, and track separate analysis scripts for each specification, which helps avoid inconsistencies or coding errors across universes. The results can then be extracted and summarized across the full set of universes to evaluate whether the conclusions are robust across reasonable analytic alternatives or depend on specific combinations of choices. The selection of analytical specifications was guided by a systematic review of prior research of TBR in ADHD (Ahmadi et al., 2020; Aldemir et al., 2018; Arns et al., 2013, 2016, 2018; Barry et al., 2009; Boxum et al., 2024; Bresnahan et al., 1999; Buyck & Wiersema, 2014a, 2014b, 2015; Chow et al., 2019; Clarke et al., 2001a, 2001b, 2002, 2011, 2020; Donoghue, 2025; Donoghue, Dominguez, et al., 2020; Dupuy et al., 2013, 2014; González-Castro et al., 2013; Halawa et al., 2017; Heinrich et al., 2014; Hermens et al., 2005; Hobbs et al., 2007; Huang et al., 2018; Janzen et al., 1995; Jarrett et al., 2020; Kitsune et al., 2015; Lansbergen et al., 2011; Lenartowicz & Loo, 2014; Liechti et al., 2013; Loo et al., 2013; Loo & Arns, 2015; Lubar, 1991; Luo et al., 2023; Markovska-Simoska & Pop-Jordanova, 2017; Martín-Brufau & Nombela-Gómez, 2017; Monastra et al., 1999; Nazari et al., 2011; Ogrim et al., 2012; Poil et al., 2014; Rezaeezadeh et al., 2020; Robertson et al., 2019; Sangal & Sangal, 2015; Shi et al., 2012; Skalski et al., 2021; Snyder et al., 2008; Woltering et al., 2012; Zhang, Johnstone, et al., 2019; Zhang, Li, et al., 2019). Following analytical specifications were investigated: resting state condition (2 levels: EO or EC), reference scheme (2 levels: average or linked mastoids), frequency band definition (2 levels: canonical frequency range [theta: 4-8 Hz; beta: 13-30 Hz] or individualized frequency bands based on IAF [theta: IAF-6 Hz to IAF-4 Hz; beta: IAF+2 Hz to 30 Hz]), handling of aperiodic signal (3 levels: uncorrected, aperiodic-adjusted, or aperiodic signal only), region of interest (6 levels: frontal (Fz), left frontal (Fp1, F3, F7), right frontal (Fp2, F4, F8), midline (Fz, Cz, Pz), central (Cz), extended central (C3, Cz, C4)), inclusion of comorbid diagnoses (2 levels: ADHD participants with comorbid diagnoses included or excluded), inclusion of subjects with medication (2 levels: ADHD participants with medication included or excluded), group contrast (2 levels: ADHD-Inattentive vs. Healthy Controls or ADHD-Combined vs. Healthy Controls), regression model (8 levels: group only; group * age; group * gender; group *IAF; group * age * gender; group * age * IAF; group * IAF * gender; group * age * gender * IAF). We use Wilkinson notation (Wilkinson & Rogers, 1973), where * indicates inclusion of both main effects and their interaction. Importantly, we focused exclusively on regression terms that included the factor group, allowing us to assess how the association between diagnostic status and TBR was moderated by age, gender, and IAF across analytical specifications. In summary, this resulted in 576 distinct model specifications (i.e., universes) per group contrast. Each universe included 8 regression terms, yielding a total of 4,608 (i.e., 576 x 8) possible effects for each group contrast (i.e. HC vs. ADHD-Inattentive and HC vs. ADHD-Combined contrasts). A full overview of the analysis tree with analytical specification and variable labeling scheme is provided in Figure 1.

As an additional robustness check, we performed a bootstrap analysis to evaluate whether unequal group sizes influenced the multiverse results. For each specification, we identified the group with the fewest participants (e.g., ADHD-Inattentive, male, unmedicated, without comorbidities) and resampled all groups to match this minimum size. The multiverse analysis was then repeated on these balanced subsamples, and the procedure was iterated 1,000 times with independent random draws. Full details and aggregated results of the bootstrap analysis are provided in the Supplement (Supplementary Material 1. 1. 4).

To complement the categorical analyses based on the diagnostic group (i.e., HC, ADHD-Inattentive, ADHD-Combined), we conducted an identical multiverse analysis using the SWAN total scores (Swanson et al., 2001) as a dimensional predictor of ADHD symptom severity in the HBN sample only (healthy controls as well as ADHD patients), as SWAN scores were not available for the validation sample. This dimensional approach tested the robustness of findings across the full spectrum of ADHD symptomatology, avoiding potential information loss from diagnostic thresholds while increasing statistical power and capturing subclinical variability. The identical set of analytical specifications and model structures were applied (i.e., 576 universes), substituting the categorical Group factor with continuous SWAN total scores and their interactions with age, gender, and IAF. Full results are provided in Supplementary Material 1. 1. 5.

#### 2.6.1. Specification curves plots

To visualize the impact of analytical specifications on the estimated effects, we constructed specification curves plots (Simonsohn et al., 2015, 2019). The specification curve displays the estimated effect size (regression coefficient) for a given regression term across all 576 analytical specifications per group contrast. This visualization provides a comprehensive overview of the consistency, direction, and statistical reliability of results across the full analytical multiverse. Separate specification curves were generated for each regression term within the HC vs. ADHD-Inattentive and HC vs. ADHD-Combined contrasts. If effect sizes remain consistent across a wide range of analytical decisions, it suggests that the findings are robust and not driven by arbitrary choices. In contrast, if results vary substantially across specifications, this indicates sensitivity to particular analytical decisions and warrants cautious interpretation. Notably, even consistently null results are informative, as they imply that no meaningful effect emerges regardless of the analytical path taken.

#### 2.6.2. Proportions plots and possibility space

To facilitate the interpretation of the results and to highlight which analytical choices most consistently produced significant effects, we summarized all 576 universes per regression term as proportion of significantly positive (Supplementary Figure 2) and negative (Supplementary Figure 3) effects. For every regression term within each ADHD subtype, we computed the proportions of significantly positive and significantly negative estimates to the total number of universes.

To assess whether the proportion of significant results within each subset of the multiverse analysis exceeded what would be expected by chance (i.e., 5%), we conducted one-sided binomial tests (i.e., binom.test()). The null hypothesis assumed that only 5% of models would yield significant results by random chance (H₀: p = 0.05), and the alternative hypothesis tested whether the observed proportion was greater than expected (H₁: p > 0.05). We computed the minimum proportion of significant results required for the binomial test to return p < 0.05 for subsets with 288 (e.g., EO and EC), 192 (e.g., uncorrected power, aperiodic-adjusted power, aperiodic signal), and 96 (e.g., 6 regions of interest) universes. The thresholds obtained from the binomial tests were approximately 7.6% (for condition, reference, frequency range, comorbidities and medication), 8.3% (for aperiodic correction), and 10.4% (for region of interest). These values were used to interpret whether observed proportions within specific analytical branches could be considered statistically robust.

When interpreting the results of a multiverse analysis, it is important to distinguish between probabilistic and possibilistic uncertainty (i.e., incertitude) (Sarma et al., 2024, 2025). Probabilistic uncertainty reflects the variability within each individual model specification, arising from sampling error and limited data. It allows for inferences about the likelihood of parameter estimates and is typically quantified through p-values, confidence intervals, or consonance curves (Fernandes et al., 2018; Sarma et al., 2024, 2025; Yang et al., 2024). In contrast, possibilistic uncertainty captures the range of results that emerge across all defensible analytic decisions within the multiverse (Sarma et al., 2024, 2025). This type of uncertainty does not imply probability. Instead, each result is treated as a valid possibility, regardless of how frequently it occurs. It is therefore essential to consider both forms of uncertainty, as they provide complementary information: probabilistic measures quantify the reliability of estimates within a given specification, while possibilistic measures reveal the robustness of conclusions across alternative specifications. To capture the range of possibilities, we developed a ShinyApp in RStudio that summarizes estimates for each regression term, visualizing the full distribution of results and highlighting significant ones through color-coding. An online version of the app is available via <u>shinyapps.io</u> (HBN: https://dstrze.shinyapps.io/HBN-sample/ and https://dstrze.shinyapps.io/hbn-sample-swan/; Validation Sample: https://dstrze.shinyapps.io/validation-sample/), and the corresponding R code is openly shared on the OSF under https://osf.io/u5yxv. Users can interactively filter model specifications to explore which analytic choices most frequently yield significant outcomes. Key plots summarizing the most important results are presented in the main text of the manuscript (Figures 5 & 7).

## 3. Results

### 3.1. Healthy Brain Network Sample

#### 3.1.1. Demographics

Table 1 and Figure 2 summarize the demographic characteristics of the Healthy Brain Network sample after all exclusions were applied, as described in the Methods section. The final sample comprised 1122 participants. Gender was compared across all three groups, that is, HC, ADHD-Combined, and ADHD-Inattentive, using a chi-square test of independence. The chi-square test revealed a significant association between group and gender (χ² = 32.60, p = 8.33e-8). Comorbid conditions and medication use were analyzed only within the ADHD sample, that is, ADHD-Combined versus ADHD-Inattentive, because these variables were not applicable to controls. Both showed significant group differences: comorbid conditions (χ² = 5.42, p = 0.019) and medication use (χ² = 38.55, p = 5.35e-10), indicating that the distributions of these variables differed between the two ADHD groups. Next, we conducted one-way ANOVAs to investigate group differences in continuous variables. The ANOVAs revealed significant group effects for age (F = 25.83, p = 1.1e-11), all SWAN subscales, including inattention (F = 237.61, p = 2.3e-86), hyperactivity (F = 279.05, p = 1.4e-98), and total scores (F = 313.25, p = 3.3e-108), as well as WISC FSIQ (F = 18.94, p = 8.6e-9), and IAF (F = 5.23, p = 0.006), indicating that the HC, ADHD-Combined and ADHD-Inattentive differed significantly across all measured variables.

**Figure 2.**
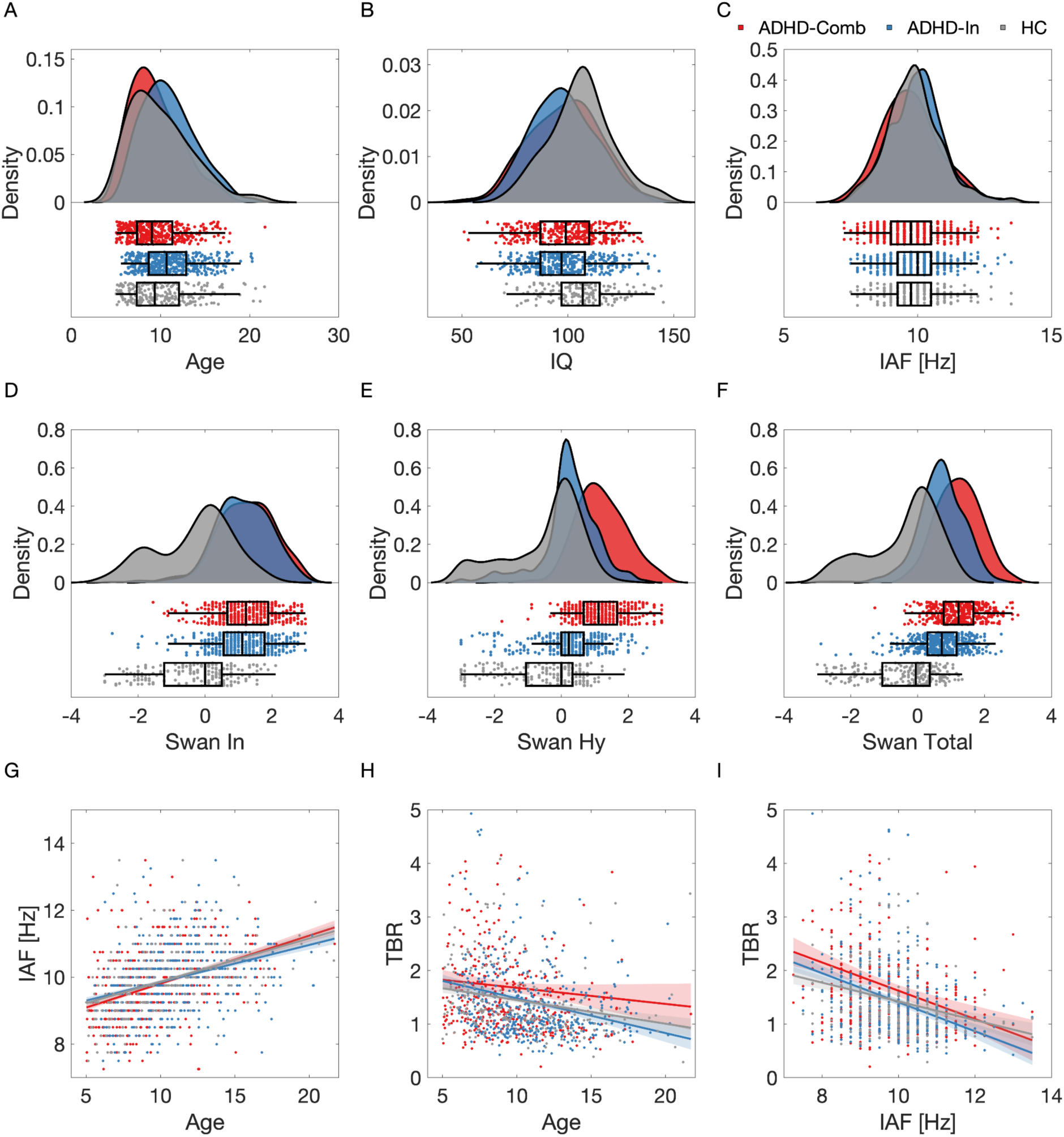
Overview of sample characteristics and variable distributions. Density plots show the distribution of (A) age, (B) IQ, (C) IAF, (D-F) and SWAN scores (Inattention, Hyperactivity, and Total) across the three groups. (G-I) Scatter plots display the relationship between (G) IAF and age, (H) between TBR and age, (I) and TBR and IAF across all participants. The regression line is shown with standard error bands. Note. IAF = Individual Alpha Frequency. TBR = Theta Beta Ratio. HC = Healthy Control.

### 3.1.2. EEG Features

To visualize the neurophysiological data, we plotted the scalp topographies of theta, beta and TBR, power spectra and aperiodic signal for HC, ADHD-Combined, and ADHD-Inattentive groups (Figure 3). The figure displays aperiodic-adjusted power during EO condition, computed using a canonical frequency range (theta: 4-8 Hz; beta: 13-30 Hz). For completeness, the corresponding 1/f-uncorrected power spectra and topographies are provided in the Supplementary Materials (1. 1. 1).

**Figure 3.**
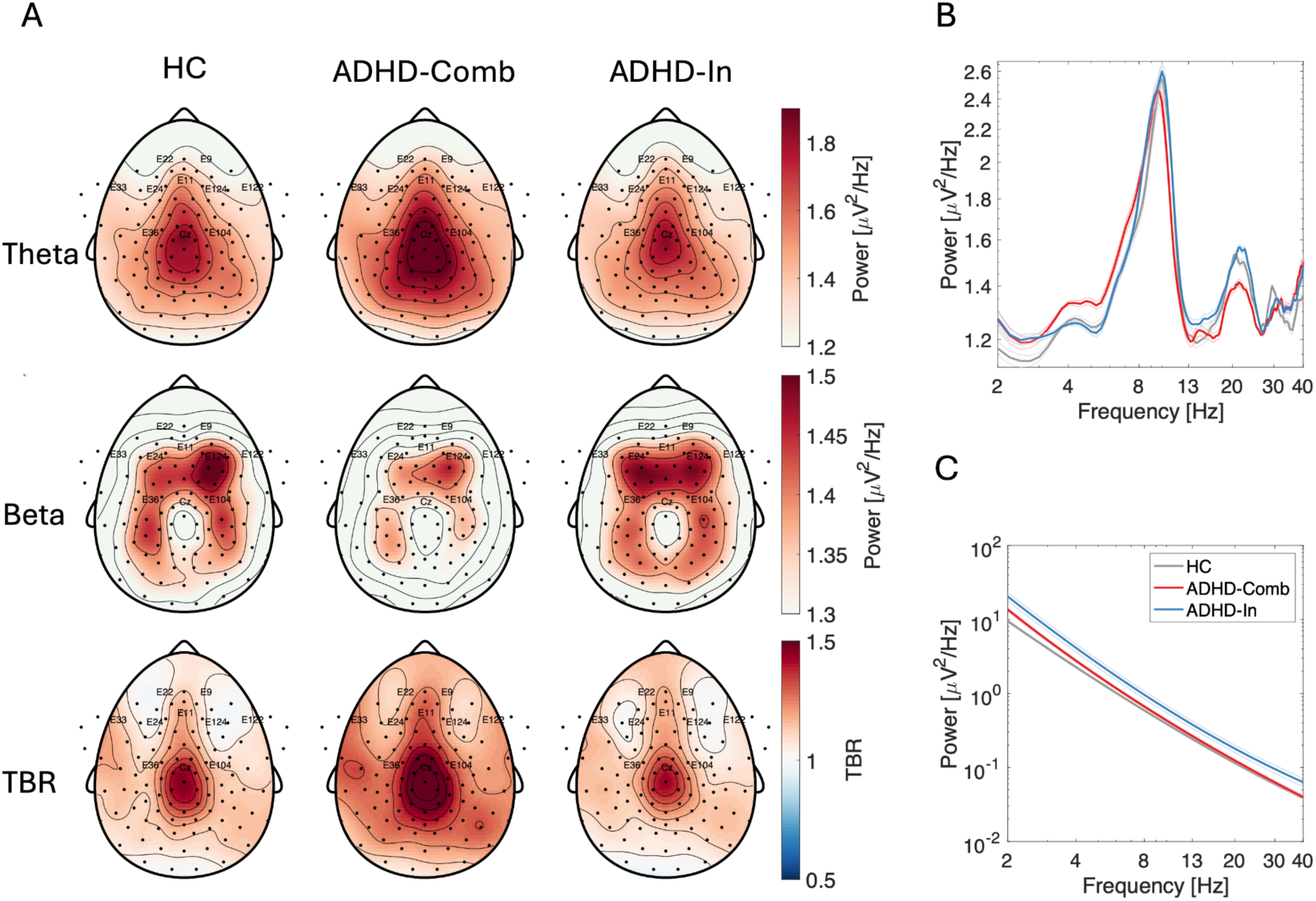
Neurophysiological data from the Healthy Brain Network sample. (A) Scalp topographies, (B) aperiodic-adjusted power spectra and (C) aperiodic signal for HC, ADHD-Combined, and ADHD-Inattentive groups. The figure displays aperiodic-adjusted power during EO condition, computed using a canonical frequency range (theta: 4-8 Hz; beta: 13-30 Hz) and TBR. The aperiodic-adjusted spectra and the reconstructed aperiodic signal are shown on a log-log scale. Electrodes labels highlighted on the topographies correspond to the six regions of interest derived from literature used across different branches of multiverse analysis. The power spectra and aperiodic signal were computed by averaging across all electrodes within each respective region of interest. Note. TBR = theta-beta ratio. HC = healthy controls. Comb = Combined. In = Inattentive.

### 3.1.3. Multiverse Analysis Results

To examine the robustness of results across all analytical specifications, we conducted a multiverse analysis using the following regression model:

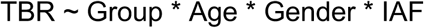

Here, Group was a factor with three levels: HC (reference), ADHD-Inattentive, and ADHD-Combined. Gender was a factor with 2 levels: male (i.e., reference) and female. Accordingly, the intercept represents healthy control males. The multiverse analysis also computed contrasts between HC and ADHD-Inattentive, as well as between HC and ADHD-Combined types. Table 2 provides a summary of the results across all multiverse specifications. In the context of the multiverse, positive universes refer to analytical specifications that yielded positive beta estimates, meaning that TBR was higher in the ADHD group than in HC. Conversely, negative universes reflect negative beta estimates, indicating lower TBR in the ADHD group relative to HC.

**Table 2.**
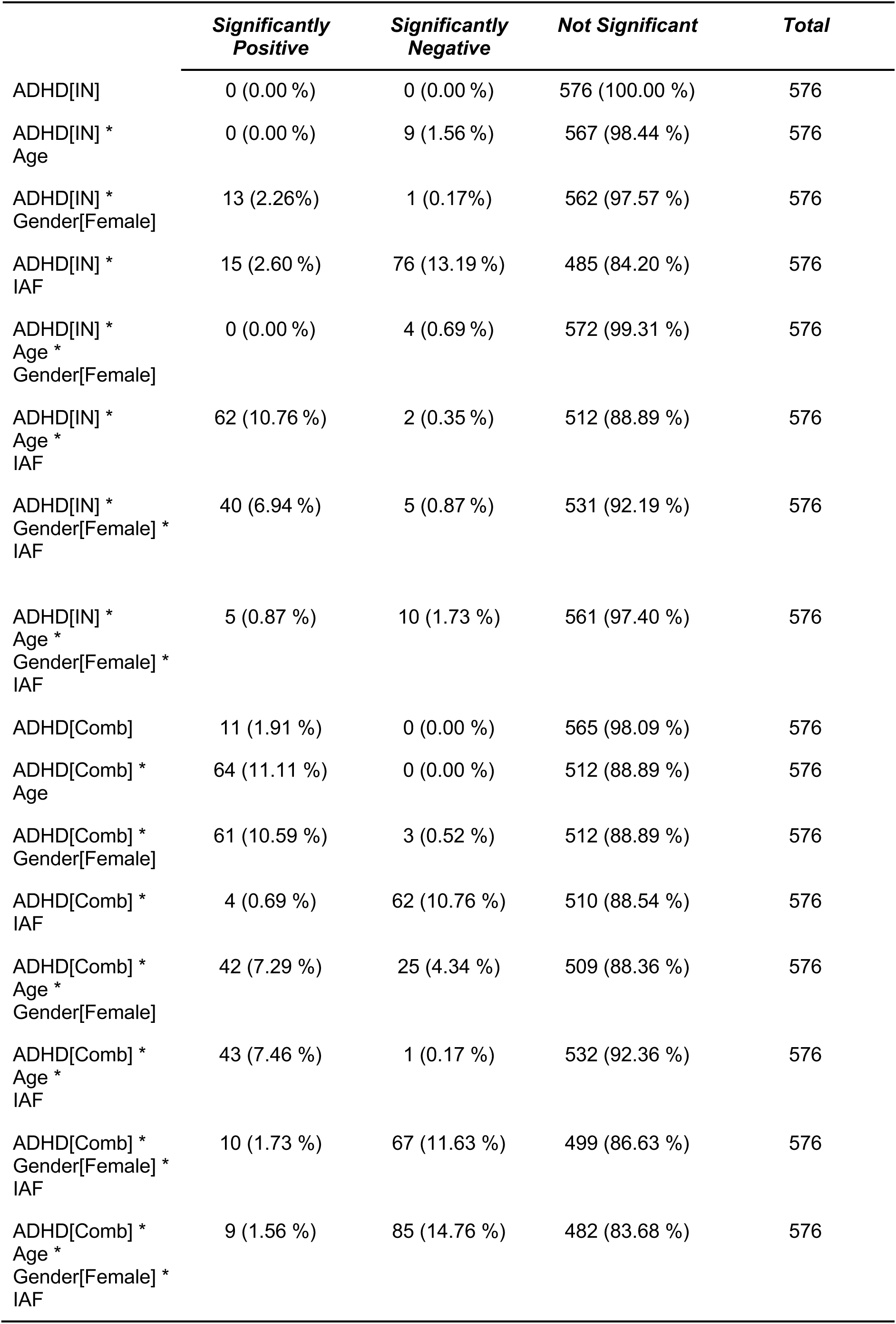
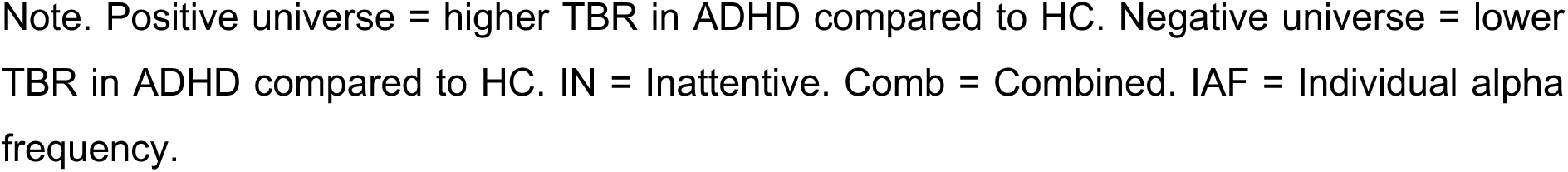
Results of the multiverse analysis in the HBN sample showing the proportion of significant effects across 576 universes.

|  | ***Significantly***<br>***Positive*** | ***Significantly***<br>***Negative*** | ***Not Significant*** | ***Total*** |
| --- | --- | --- | --- | --- |
| ADHD[IN] | 0 (0.00 %) | 0 (0.00 %) | 576 (100.00 %) | 576 |
| ADHD[IN] *<br>Age | 0 (0.00 %) | 9 (1.56 %) | 567 (98.44 %) | 576 |
| ADHD[IN] *<br>Gender[Female] | 13 (2.26%) | 1 (0.17%) | 562 (97.57 %) | 576 |
| ADHD[IN] *<br>IAF | 15 (2.60 %) | 76 (13.19 %) | 485 (84.20 %) | 576 |
| ADHD[IN] *<br>Age *<br>Gender[Female] | 0 (0.00 %) | 4 (0.69 %) | 572 (99.31 %) | 576 |
| ADHD[IN] *<br>Age *<br>IAF | 62 (10.76 %) | 2 (0.35 %) | 512 (88.89 %) | 576 |
| ADHD[IN] *<br>Gender[Female] *<br>IAF | 40 (6.94 %) | 5 (0.87 %) | 531 (92.19 %) | 576 |
| ADHD[IN] *<br>Age *<br>Gender[Female] *<br>IAF | 5 (0.87 %) | 10 (1.73 %) | 561 (97.40 %) | 576 |
| ADHD[Comb] | 11 (1.91 %) | 0 (0.00 %) | 565 (98.09 %) | 576 |
| ADHD[Comb] *<br>Age | 64 (11.11 %) | 0 (0.00 %) | 512 (88.89 %) | 576 |
| ADHD[Comb] *<br>Gender[Female] | 61 (10.59 %) | 3 (0.52 %) | 512 (88.89 %) | 576 |
| ADHD[Comb] *<br>IAF | 4 (0.69 %) | 62 (10.76 %) | 510 (88.54 %) | 576 |
| ADHD[Comb] *<br>Age *<br>Gender[Female] | 42 (7.29 %) | 25 (4.34 %) | 509 (88.36 %) | 576 |
| ADHD[Comb] *<br>Age *<br>IAF | 43 (7.46 %) | 1 (0.17 %) | 532 (92.36 %) | 576 |
| ADHD[Comb] *<br>Gender[Female] *<br>IAF | 10 (1.73 %) | 67 (11.63 %) | 499 (86.63 %) | 576 |
| ADHD[Comb] *<br>Age *<br>Gender[Female] *<br>IAF | 9 (1.56 %) | 85 (14.76 %) | 482 (83.68 %) | 576 |

Note. Positive universe = higher TBR in ADHD compared to HC. Negative universe = lower TBR in ADHD compared to HC. IN = Inattentive. Comb = Combined. IAF = Individual alpha frequency.

#### 3.1.3.1. Healthy controls vs. ADHD-Inattentive comparison

The multiverse analysis revealed 0 significantly positive (i.e., higher TBR in ADHD-Inattentive compared to HC) and 0 significantly negative (i.e., lower TBR in ADHD-Inattentive compared to HC) universes out of 576 for the main effect of ADHD-Inattentive, indicating that TBR did not differ between ADHD-IN males compared to healthy control males in any specification (Figure 4).

**Figure 4.**
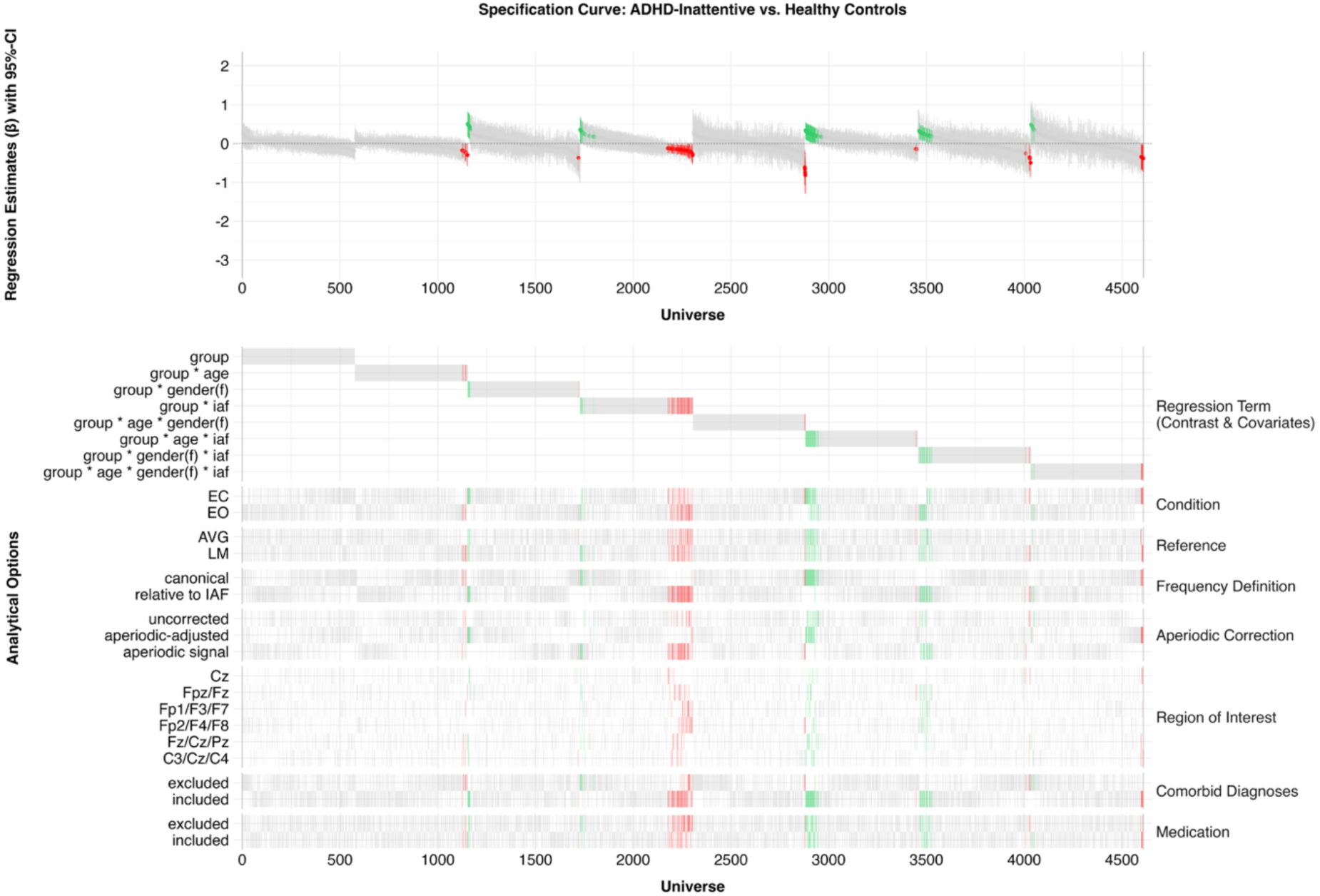
Specification curve representing all universes for the healthy control ADHD-Inattentive contrast. The top panel shows regression estimates for each specification, sorted by regression estimate, with 95% confidence intervals. Statistically significant positive estimates are shown in green, negative estimates in red, and non-significant estimates in gray. The bottom panel indicates which analytical choices were associated with significantly positive (i.e., higher TBR for ADHD compared to HC) or negative (i.e., lower TBR for ADHD compared to HC) effects across specifications. Note. CI = confidence interval. f = female. EC = eyes closed. EO = eyes open. AVG = average reference. LM = linked mastoid reference. canonical = canonical frequency range. relative to IAF = bandwidth relative to individual alpha frequency.

Notably, the most frequently observed effects in the ADHD-Inattentive vs. healthy control comparisons involved the IAF, specifically the interactions ADHD-Inattentive * IAF, ADHD-Inattentive * Age * IAF, and ADHD-Inattentive * Gender * IAF. As illustrated in the Figure 4, the negative ADHD-Inattentive * IAF effect emerged almost exclusively when using frequency bands relative to IAF and was most frequently observed for the aperiodic signal and when children with comorbidities were included. To further explore the commonalities across universes, we used the ShinyApp to interactively filter the analytical space. When restricting the multiverse to the models with frequency bands relative to IAF, aperiodic signal as spectral measure and including subjects with comorbidities, all 48 remaining universes (i.e., across 2 resting-state conditions × 2 references × 6 ROIs × 2 medication options) yielded negative estimates, with 46 of them reaching statistical significance (Figure 5 A). In contrast, the positive ADHD-Inattentive * Age * IAF effect was almost exclusively found when TBR was computed using canonical frequency bands and aperiodic-adjusted power, and in analyses that included participants with comorbid diagnoses (37 significant universes out of 48 possibilities; Figure 5 B). Finally, the positive ADHD-Inattentive * Gender * IAF effect was observed exclusively when using frequency bands relative to IAF, primarily in the aperiodic signal, and again in analyses that included children with comorbid diagnoses (28 significant universes out of 48 possibilities; Figure 5 C).

**Figure 5.**
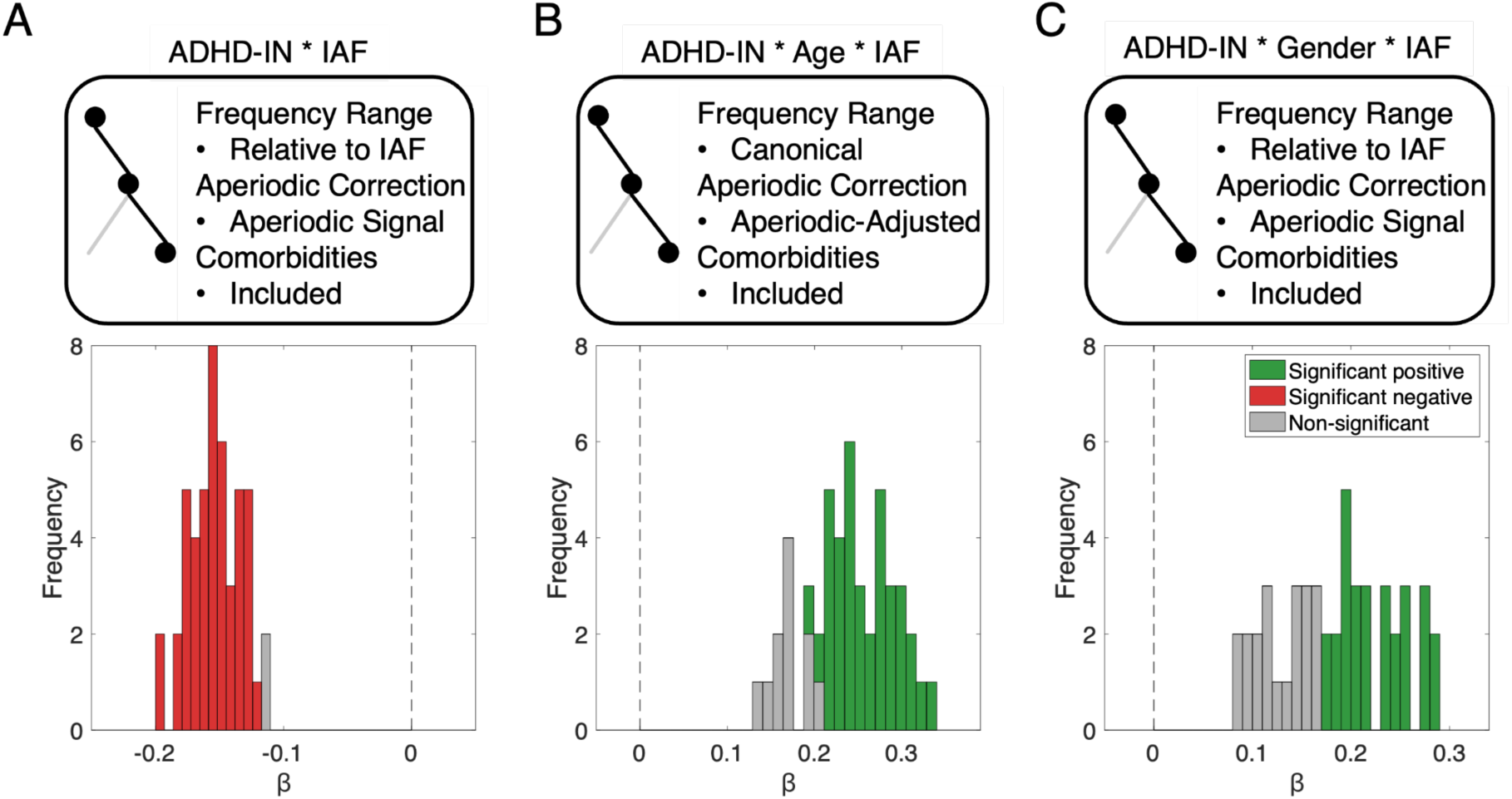
Possibility space of regression estimates from the subset of analytical choices yielding the highest number of significant results. (A) ADHD-Inattentive * IAF interaction: Significant effects (46 significant out of 48 possibilities) were predominantly observed when using frequency bands relative to IAF, the aperiodic signal, and including children with comorbid diagnoses. (B) ADHD-Inattentive * Age * IAF interaction: This positive effect emerged almost exclusively when TBR was computed using canonical frequency bands, aperiodic-adjusted power, and analyses included participants with comorbidities (37 out of 48 significant). (C) ADHD-Inattentive * Gender * IAF interaction: The effect was found exclusively under frequency bands relative to IAF, primarily in the aperiodic signal, and when children with comorbid diagnoses were included (28 out of 48 significant). Note. The significant positive estimates are highlighted green, significant negative in red and non-significant in grey.

#### 3.1.3.2. Healthy controls vs. ADHD-Combined comparison

Next, we compared the ADHD-Combined group to healthy controls across all analytical specifications. The multiverse analysis revealed 11 significantly positive (i.e., higher TBR in ADHD-Combined compared to HC) and 0 significantly negative (i.e., lower TBR in ADHD-Combined compared to HC) universes out of 576 for the main effect of ADHD-Combined, indicating that TBR was higher in ADHD-Combined males compared to healthy control males in 11 specifications (Figure 6).

**Figure 6.**
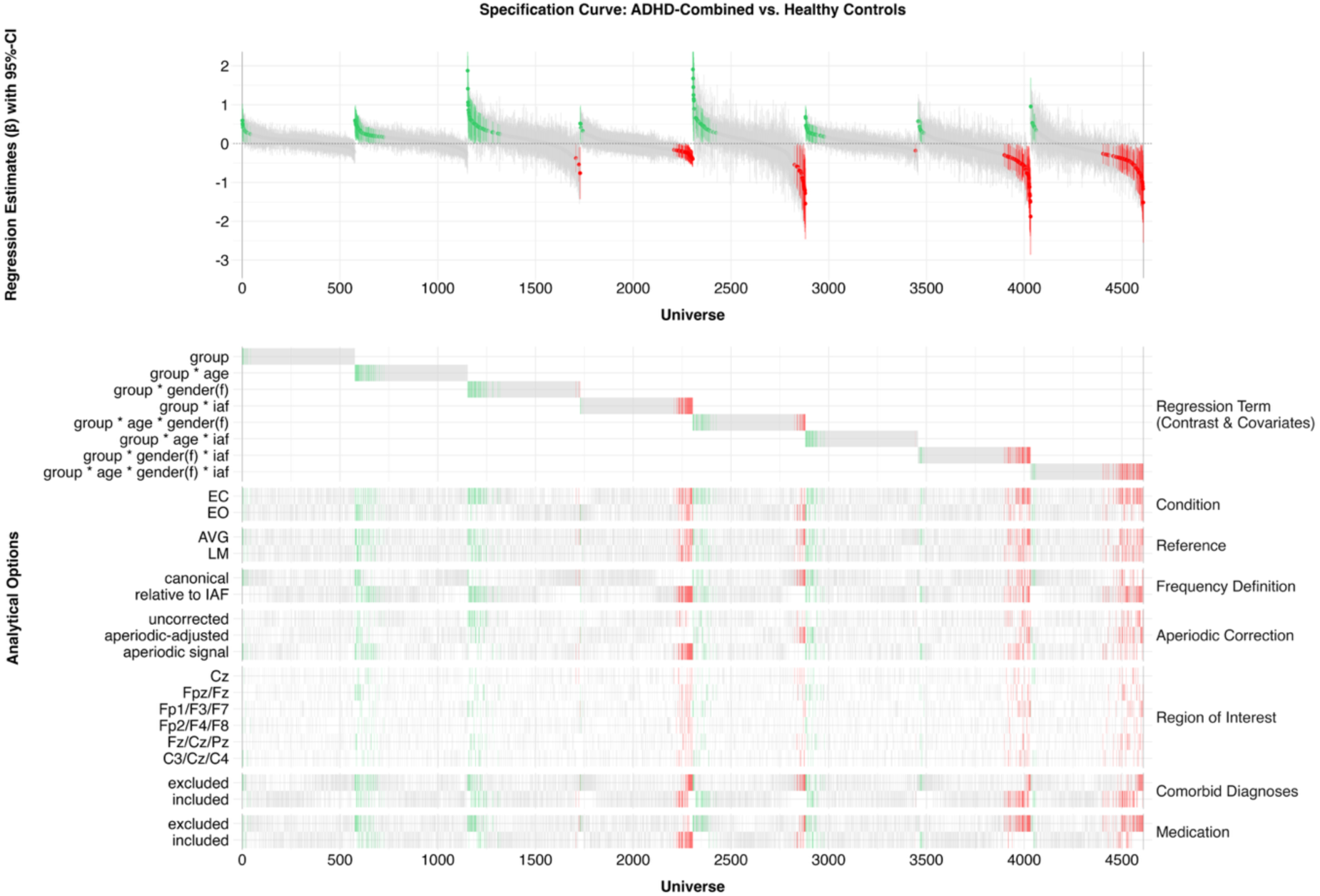
Specification curve representing all universes for the healthy control ADHD-Combined contrast. The top panel shows regression estimates for each specification, sorted by regression estimate, with 95% confidence intervals. Statistically significant positive estimates are shown in green, negative estimates in red, and non-significant estimates in gray. The bottom panel indicates which analytical choices were associated with significantly positive (i.e., higher TBR for ADHD compared to HC) or negative (i.e., lower TBR for ADHD compared to HC) effects across specifications. Note. CI = confidence interval. f = female. EC = eyes closed. EO = eyes open. AVG = average reference. LM = linked mastoid reference. canonical = canonical frequency range. relative to IAF = bandwidth relative to individual alpha frequency.

Notably, the most frequently observed effects in the ADHD-Combined vs. healthy control comparisons involved interactions with Age, Gender, and IAF. As illustrated in Figure 6, the positive ADHD-Combined * Age effect emerged mostly when using frequency bands relative to IAF and was most frequently observed in the aperiodic signal (24 significant universes out of 96 possibilities; Figure 7 A). Similarly, the negative ADHD-Combined * IAF effect occurred almost exclusively when using frequency bands relative to IAF, was predominantly observed in the aperiodic signal, and emerged mainly in analyses that included children on medication (47 significant universes out of 48 possibilities; Figure 7 B). The positive ADHD-Combined * Gender effect appeared almost exclusively in the EC condition, primarily when using frequency bands relative to IAF, and was most often found in 1/f-uncorrected power (27 significant universes out of 48 possibilities; Figure 7 C). The positive ADHD-Combined * Age * IAF effect showed no clear pattern across specifications and was relatively evenly distributed. Finally, the three interactions including gender (i.e., positive ADHD-Combined * Age * Gender, ADHD-Combined * Gender * IAF and ADHD-Combined * Age * Gender * IAF) appeared most frequently in the EC condition, almost exclusively when children with comorbid diagnoses were included, but those on medication were excluded (26 significant universes out of 72 possibilities, 35 significant universes out of out of 72 possibilities and 31 significant universes out of out of 72 possibilities, respectively; Figure 7 D-F).

**Figure 7.**
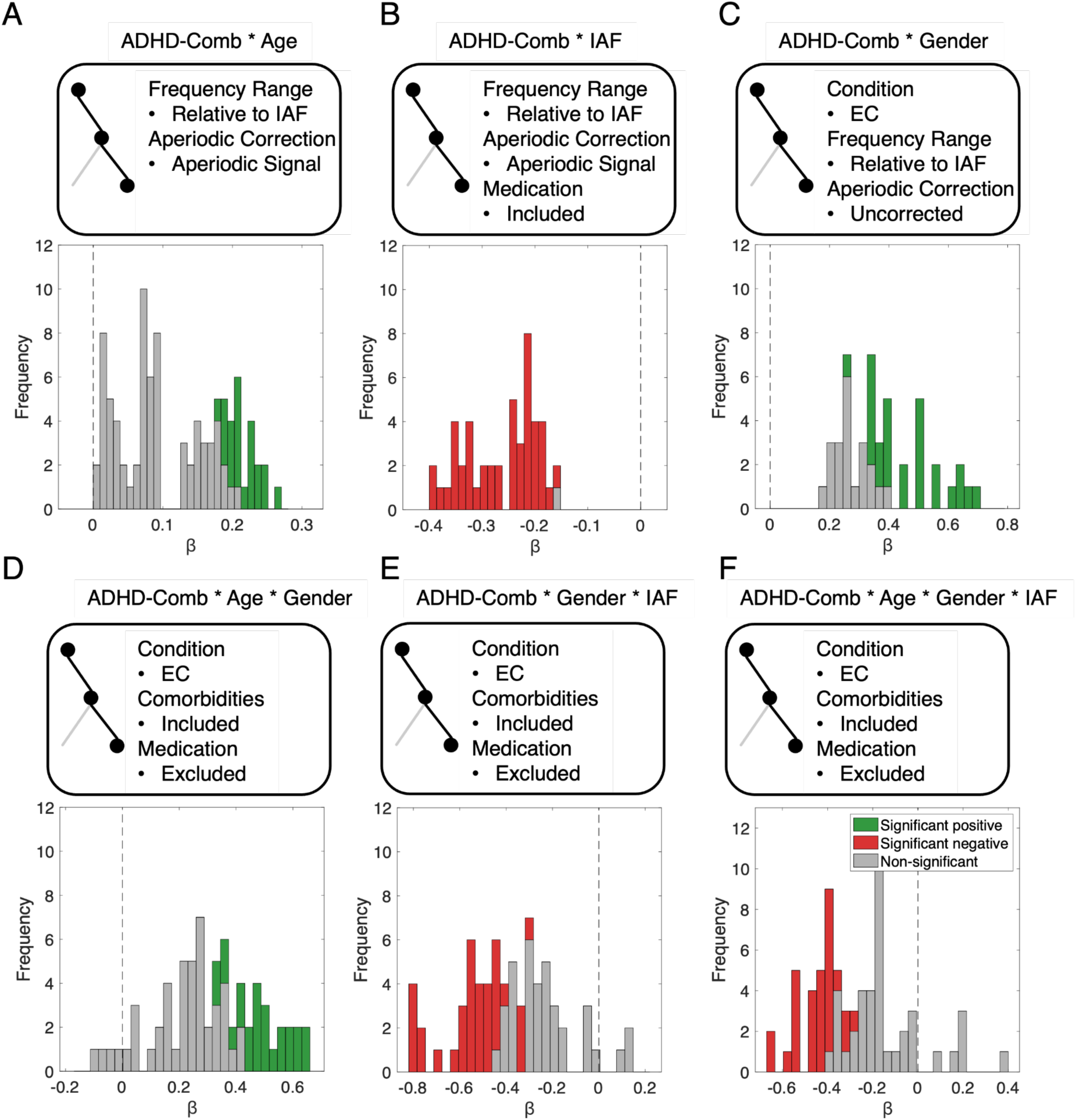
Possibility space of regression estimates from the subset of analytical choices yielding the highest number of significant results in the ADHD-Combined vs. healthy control comparisons. (A) ADHD-Combined * Age interaction: Significant effects (24 out of 96) were primarily observed when using frequency bands relative to IAF and the aperiodic signal. (B) ADHD-Combined * IAF interaction: A negative effect emerged almost exclusively under frequency bands relative to IAF, predominantly in the aperiodic signal, and in analyses including children on medication (47 out of 48 significant). (C) ADHD-Combined * Gender interaction: This positive effect appeared almost exclusively in the EC condition, with frequency bands relative to IAF and 1/f-uncorrected power (27 out of 48 significant). (D) ADHD-Combined * Age * Gender, (E) ADHD-Combined * Gender * IAF and (F) ADHD-Combined * Age * Gender * IAF interactions: These effects appeared mainly in the EC condition, almost exclusively when comorbidities were included and medication excluded (26, 35 and 31 out of 72 significant, respectively). Note. Significant positive estimates are highlighted green, significant negative in red and non-significant estimates are shown in grey.

### 3.1.4. Effect sizes across analytical subspaces

To complement the regression coefficients from the multiverse models, we additionally examined descriptive standardized effect sizes across representative analytical subspaces. Analytical paths were grouped according to frequency band definition (IAF-relative vs. canonical) and spectral representation (aperiodic signal, 1/f-uncorrected power, and aperiodic-adjusted power). Within each subspace, Cohen’s d was computed for theta power, beta power, and TBR for both the HC vs. ADHD-Inattentive and HC vs. ADHD-Combined comparisons. To visualize the distribution of effect sizes across the analytical space, violin plots were constructed with each data point representing the Cohen’s d value of a single analytical specification (Figure 8). Across all subspaces and outcome measures, Cohen’s d values were small for both comparisons, including subspaces in which interaction effects frequently reached statistical significance in the multiverse analysis. This pattern indicates that even where the multiverse revealed reliable significant effects, the underlying group differences in theta power, beta power, and TBR remained small in magnitude. These findings support the interpretation that the significant interactions observed across analytical specifications are driven by subtle moderation effects and analytical choices rather than large, robust group differences in neural activity.

**Figure 8.**
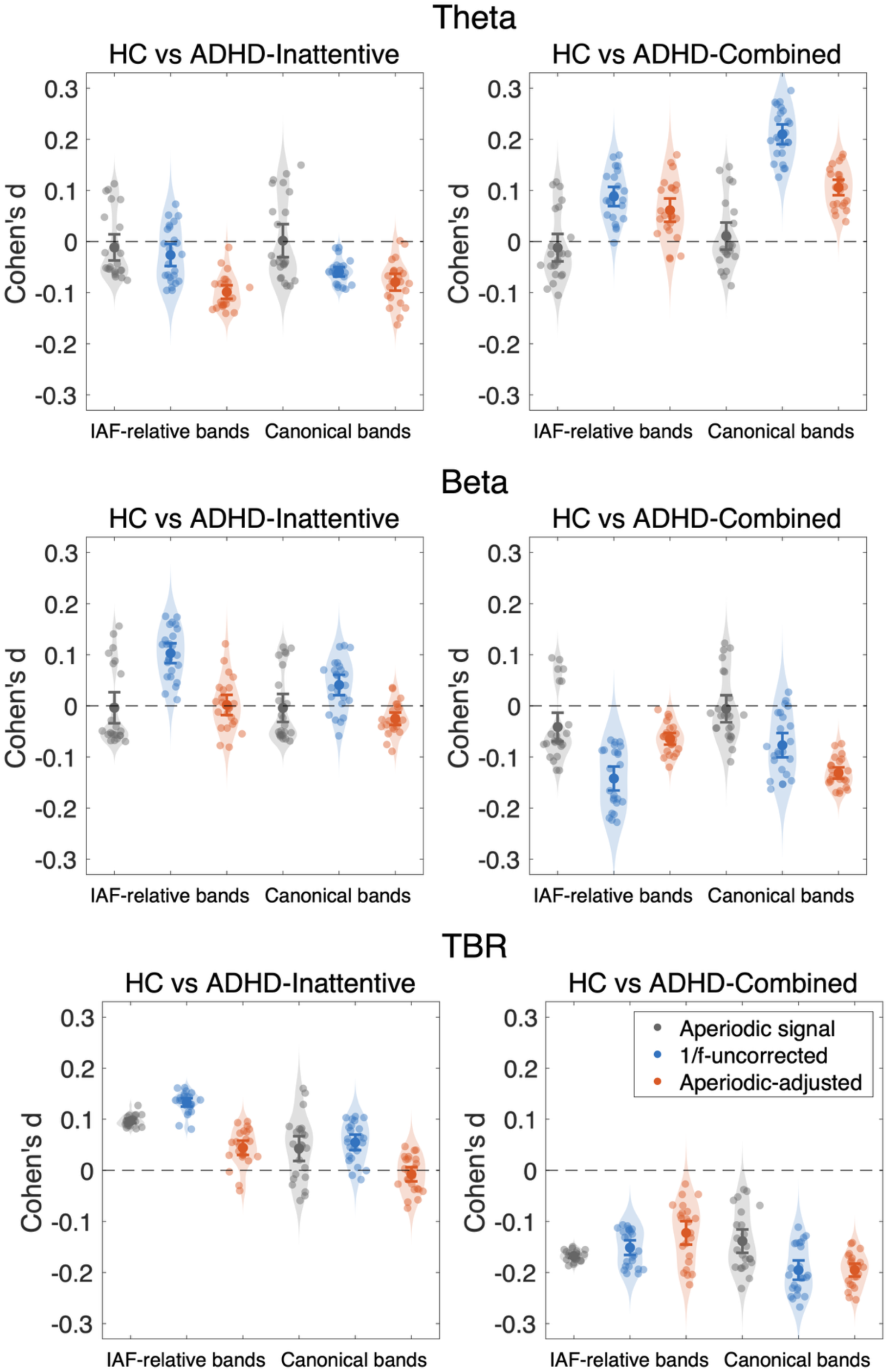
Distribution of effect sizes across analytical subspaces. Violin plots display the distribution of Cohen’s d values across analytical paths for the HC versus ADHD-Inattentive comparison and the HC versus ADHD-Combined comparison. Subspaces were defined by crossing frequency band definition (IAF-relative vs canonical frequency bands) with spectral representation (aperiodic signal, 1/f-uncorrected power, and aperiodic-adjusted power). Individual dots represent effect sizes from single analytical paths, while the error bars indicate mean effect sizes and 95% confidence intervals within each subspace. Despite frequent significant interaction effects observed in some multiverse subspaces, the corresponding effect sizes remained small across analyses. Positive values indicate higher values in healthy controls relative to ADHD groups.

### 3.1.5. Robustness analyses

To assess whether our multiverse findings were influenced by unequal group sizes across predictors, we conducted a bootstrap analysis in which all groups were resampled to the size of the smallest subgroup within each specification. For example, we identified the group with the lowest number of participants (e.g., ADHD-Inattentive, female, medication and comorbidities excluded) and randomly resampled all groups to this minimum size. This procedure was repeated 1,000 times with independent random draws. The results closely mirrored the main multiverse findings: the same interaction patterns involving IAF, age, and gender consistently emerged, and the frequency and direction of significant effects were relatively stable across iterations, as illustrated by low standard deviations. These results indicate that the observed patterns are robust and not a bias introduced by group size imbalance (see Supplementary Material 1. 1. 4).

To examine whether our results depended on categorical diagnosis definitions, we repeated the multiverse analysis using dimensional SWAN scores as continuous predictors. The dimensional multiverse analysis yielded results that were highly similar to the categorical findings (Supplementary Material 1. 1. 5). The main effect of SWAN showed no significantly positive universes (i.e., higher TBR in ADHD-Combined compared to HC) and only 3.65% significantly negative universes (i.e., lower TBR in ADHD-Combined compared to HC) across all specifications. Consistent with the categorical approach, the most frequent significant interactions involved age, gender, and IAF: SWAN * Gender interactions were significantly positive in 11.28% of specifications, SWAN * IAF interactions yielded significantly negative effects in 23.26% of specifications, SWAN * Gender * IAF showed significantly positive effects in 13.71% of specifications, and the four-way SWAN * Age * Gender * IAF interaction demonstrated significantly negative effects in 11.63% of cases. The convergence between categorical and dimensional approaches validates the robustness of the observed TBR effects in ADHD, demonstrating that these associations are consistent regardless of whether ADHD is operationalized as discrete diagnostic categories or as a continuous symptom dimension. Detailed analyses are presented in Supplementary Material 1. 1. 5.

## 4. Discussion

The present study examined the robustness of theta-beta ratio (TBR) differences between ADHD subtypes and healthy controls using a multiverse analysis across two independent samples. By systematically varying reasonable analytical parameters, including resting-state condition, frequency band definitions, aperiodic correction, reference scheme, region of interest, and inclusion/exclusion of comorbidities and medication, we quantified both the likelihood of detecting ADHD-TBR associations and the specific analytic conditions under which they arise. This approach directly addresses long-standing inconsistencies in EEG biomarker research and reveals why previous findings on TBR in ADHD have been so heterogeneous. Our results show that TBR differences are highly contingent on analytic choices. Robust effects did not emerge as simple main effects of diagnosis but rather as higher-order interactions with individual alpha frequency (IAF), age, and gender, particularly in universes without 1/f correction and those using IAF-relative frequency bands. Importantly, these interaction patterns were strikingly similar across the two independent datasets and held for both categorical and dimensional definitions of ADHD. Below, we first discuss the role of covariates in shaping TBR, then place these findings in a developmental context, and finally consider their clinical implications.

### No Reliable Main Effects of Diagnosis on TBR

Across the 576 universes in the HBN sample and the 144 universes in the validation sample, we found no consistent TBR differences between healthy controls and either ADHD subtypes. For the HC-ADHD-Inattentive comparison, positive universes (indicating higher TBR in ADHD than in HC) and negative universes (indicating lower TBR in ADHD compared to HC) were entirely absent in the HBN sample (0% positive, 0% negative) and similarly rare in the validation sample (1.39% positive, 7.64% negative). The same pattern emerged for ADHD-Combined, with only minimal proportions of positive or negative universes (HBN: 1.91% positive, 0% negative; validation: 0.69% positive, 1.39% negative). Importantly, the results from the categorical approach were corroborated by the dimensional analyses using SWAN scores, indicating convergence across methodological frameworks (Supplementary Material 1. 1. 5). These null results align with a growing body of work challenging the diagnostic value of TBR (Arns et al., 2013, 2016, 2018; Boxum et al., 2024; van Dijk et al., 2020). Earlier reports of large group differences have increasingly been attributed to methodological limitations, sample imbalances, and analytic flexibility, while more recent studies have repeatedly failed to identify reliable ADHD-related TBR effects. Our multiverse results thus converge with this broader literature, providing further evidence that TBR lacks the reliability and discriminative validity required for clinical utility. Beyond methodological convergence across analytical frameworks, the consistency of results across two datasets differing substantially in EEG recording systems and impedance thresholds further strengthens the generalizability of these null findings, suggesting they are unlikely to reflect idiosyncrasies of a specific acquisition protocol. Taken together, these findings demonstrate that, regardless of whether ADHD is operationalized as discrete diagnostic categories or as a continuous symptom dimension, TBR alone shows insufficient diagnostic reliability for differentiating healthy controls from individuals with ADHD.

### The role of IAF and 1/f on TBR differences

Although we found no consistent main effects of ADHD diagnosis on TBR, our multiverse analyses revealed recurring interaction patterns involving IAF, age, and gender. These interactions were highly dependent on analytic decisions, most notably, the choice between canonical versus IAF-relative frequency bands and whether oscillatory and aperiodic components were separated. Across both samples, significant effects occurred most frequently in models using IAF-relative bands combined with either the aperiodic slope or uncorrected 1/f signal. In contrast, when canonical frequency bands or aperiodic-adjusted power were used, these effects largely disappeared. These results suggest that apparent TBR differences may reflect properties of the aperiodic background signal interacting with individual variability in IAF rather than true oscillatory theta or beta activity. This interpretation is consistent with previous work showing that electrophysiological frequency band ratio measures can conflate periodic and aperiodic neural activity, such that apparent changes in theta/beta or other band ratios may partly reflect changes in the aperiodic spectral component rather than narrowband oscillatory activity (Donoghue, 2025; Donoghue, Dominguez, et al., 2020).

Two mechanisms are likely to contribute to these effects. First, subtle differences in aperiodic slope disproportionately affect low frequencies, where the 1/f structure inflates amplitude and can mimic elevated theta power. Second, differences in IAF shift the frequency bands such that individuals with slower IAF have theta bands defined at lower frequencies (e.g., individual with slow IAF = 8 Hz has theta band defined at 2-4 Hz), which are more strongly influenced by the aperiodic signal. It artificially elevates TBR, whereas faster IAF is associated with lower TBR estimates. Importantly, both mechanisms can operate simultaneously and amplify each other. Even if group differences in IAF or aperiodic slope alone are insufficient to distinguish healthy controls from ADHD subtypes, their combined influence can produce statistically significant interaction effects. For example, when fixed canonical bands were used, IAF correlates only modestly with 1/f-uncorrected (r = –0.17, p = 1.28e-7) or aperiodic-adjusted power (r = –0.18, p = 1.65e-9). However, when frequency bands are defined relative to IAF, the correlation between IAF and 1/f-uncorrected power becomes markedly stronger (r = –0.70, p = 2e-308), and even reverses direction for aperiodic-adjusted power (r = 0.11, p = 1.47e-4), demonstrating how the interaction between subtle changes in 1/f-slope and IAF might greatly affect TBR estimates. Furthermore, our data show overall group differences in IAF (Table 1), although post-hoc comparisons indicate no significant differences between healthy controls and either ADHD-Combined (t(655) = 1.55, p = 0.122) or ADHD-Inattentive (t(691) = –1.09, p = 0.275) subgroups. The only statistically significant difference was a higher IAF in ADHD-Inattentive compared to ADHD-Combined (t(892) = –3.21, p = 0.001). Thus, while neither IAF nor aperiodic slope significantly differentiates HC from ADHD subtypes, their interaction magnified by analytic choices such as IAF-relative band definitions can produce group-by-IAF interactions in TBR.

An additional explanation previously proposed in the literature is that individuals with slow IAF may appear to have elevated theta power because the lower alpha range overlaps with canonical theta frequencies. In such cases, alpha power may leak into the theta band when fixed frequency ranges are used (Boxum et al., 2024; Klimesch, 1999). This interpretation is consistent with findings showing that increased TBR in boys with ADHD was replicated only when canonical frequency bands were applied but disappeared once IAF-relative frequency ranges were used (Lansbergen et al., 2011). However, our results do not support the idea that low IAF produces artificially elevated theta power through alpha leakage. We observed no such effect in either the 1/f-uncorrected power or in the aperiodic-adjusted power analyses, when canonical frequency bands were used. Instead, the pattern of results suggests that IAF-related variation in TBR is driven primarily by differences in aperiodic spectral slope, as discussed above, rather than by overlap between alpha and theta frequencies.

IAF also has developmental and clinical relevance that further contextualizes these findings. With maturation, IAF typically increases, shifting theta bands closer toward the canonical 4-8 Hz range (Tröndle et al., 2022a). As a result, theta power derived from 1/f-uncorrected power tends to decrease with age, since the influence of the aperiodic component diminishes at higher frequencies. These dynamics may account for the recurring Age * IAF interactions observed in our multiverse analyses, as both oscillatory and aperiodic contributions vary systematically across the lifespan (Hill et al., 2022; McSweeney et al., 2023; Merkin et al., 2023; Stanyard et al., 2024; Tröndle et al., 2021, 2022a). These developmental considerations highlight the importance of explicitly modeling the aperiodic component when studying neural oscillations over lifespan. Oscillatory and aperiodic activity represent distinct neurophysiological phenomena, yet they can easily contaminate each other if not separated. The functional meaning of the aperiodic component remains debated: it may reflect neural processes related to excitatory-inhibitory balance (Donoghue, Haller, et al., 2020; Gao et al., 2017), but it may also partly capture measurement artifacts without direct biological relevance (Schmidt et al., 2025; Tröndle & Langer, 2026a). Importantly, recent evidence suggests that 1/f intercept and slopes can be systematically affected by various artifacts (i.e., heart, eye, muscle). If one group contributes systematically more artifacts to the EEG signal, for example, due to increased movement in participants with ADHD (Dziemian et al., 2022), this can artificially steepen the estimated 1/f slope and elevate low-frequency power. Such differences in artifact susceptibility make it difficult to determine whether observed TBR effects reflect genuine neurophysiological variation or shifts in spectral composition caused by noise. Future work should therefore apply methods that separate oscillatory from aperiodic activity across development and clinical groups, while ideally also measuring movement artifacts (e.g., via EMG), in order to clarify whether apparent TBR differences reflect true neural alterations or shifts in spectral composition and artifact susceptibility.

While TBR appears to lack diagnostic reliability, emerging evidence suggests that other EEG features may be more informative for clinical decision-making. In particular, IAF has been proposed as a marker for predicting treatment response, with IAF-based stratification improving remission rates when selecting between methylphenidate and neurofeedback interventions (Boxum et al., 2024; Krepel et al., 2020; Voetterl et al., 2023). Lower frontal IAF has also been reported in adolescent non-responders (Arns et al., 2018), potentially reflecting a disruption or delay in the normative developmental increase of IAF. However, our findings indicate that IAF alone is insufficient. Accurate interpretation of spectral biomarkers must also account for individual differences in the aperiodic 1/f slope, which strongly influence power estimates and their apparent associations with behavioral or clinical measures. Together, our results indicate that TBR is not a reliable biomarker of ADHD. Instead, TBR effects are strongly shaped by analytic decisions that alter how oscillatory and aperiodic components are represented. Such analytic dependence may help explain the long-standing inconsistency in TBR findings across the ADHD literature, as unaccounted differences in IAF and aperiodic slope can systematically bias power estimates, particularly when frequency bands are defined relative to IAF or when aperiodic activity is not adequately separated from oscillatory power.

### Conclusion

In this study, we used a multiverse analysis across two large EEG datasets to systematically examine the robustness of TBR as an ADHD biomarker. Across hundreds of plausible analytical specifications, we found no consistent evidence that TBR differentiates individuals with ADHD from healthy controls. Instead, TBR estimates varied substantially depending on analytic choices, most notably whether frequency bands were defined relative to IAF and how oscillatory versus aperiodic components were handled. Our findings suggest that individual differences in IAF and the aperiodic 1/f slope strongly influence TBR values and can create the appearance of group differences even when underlying oscillatory activity is similar. Although TBR itself does not provide a reliable diagnostic marker, individualized spectral features, particularly IAF in combination with the aperiodic slope, may offer more promising and clinically informative alternatives. Future work should therefore prioritize analytic frameworks that explicitly separate oscillatory and aperiodic activity and incorporate IAF when developing EEG-based biomarkers for ADHD.

## Data Availability

All data produced are available online at https://fcon_1000.projects.nitrc.org/indi/cmi_healthy_brain_network/

