## Supplement for "Theta-Beta Ratio in Attention Deficit Hyperactivity Disorder: A Multiverse Analysis"

### 1. Supplementary Material

#### 1.1. Healthy Brain Network Sample

##### 1.1.1. 1/f-uncorrected power

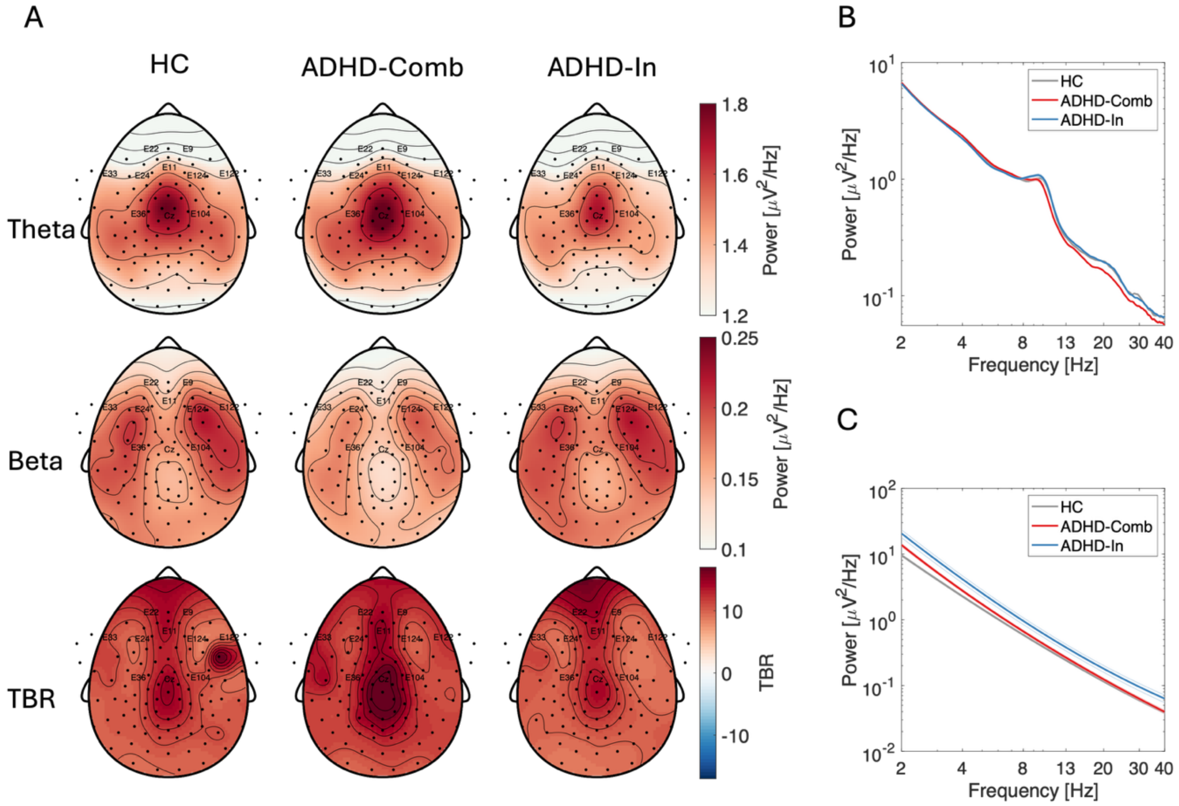


Supplementary Figure 1. Neurophysiological data from the Healthy Brain Network sample. (A) Scalp topographies, (B) 1/f-uncorrected power spectra and (C) aperiodic signal for HC, ADHD-Combined, and ADHD-Inattentive groups. The figure displays 1/f-uncorrected power during EO condition, computed using a fixed frequency range (theta: 4-8 Hz; beta: 13-30 Hz) and TBR. The 1/f-uncorrected spectra and the reconstructed aperiodic signal are shown on a log-log scale. Electrodes highlighted on the topographies correspond to the six regions of interest derived from literature used across different branches of multiverse analysis. The power spectra and aperiodic signal were computed by averaging across all electrodes within each respective region of interest. Note. HC = healthy controls. Comb = Combined. In = Inattentive.

##### 1.1.2. Proportion Plots Across All Analytical Specifications

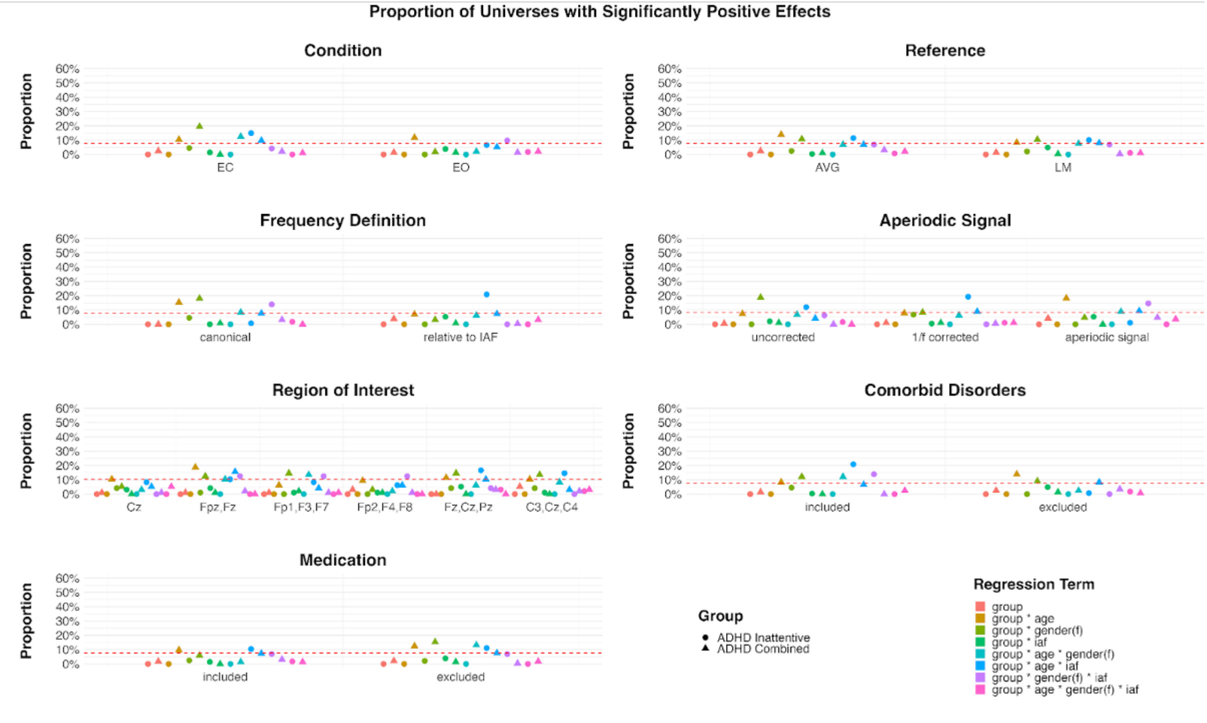


Supplementary Figure 2. Proportion of specifications showing a significantly positive effect by regression term and ADHD subtype across all analytical specifications. Each level of Condition, Reference, Frequency Definition, Comorbid Disorder, and Medication includes 288 analytical universes. The Aperiodic Signal factor includes 192 universes per level, and the Region of Interest includes 96 universes per level. The red line in the plot indicates the exact proportion of significant results required for the binomial test to reach statistical significance ($p < .05$) for each subset size.

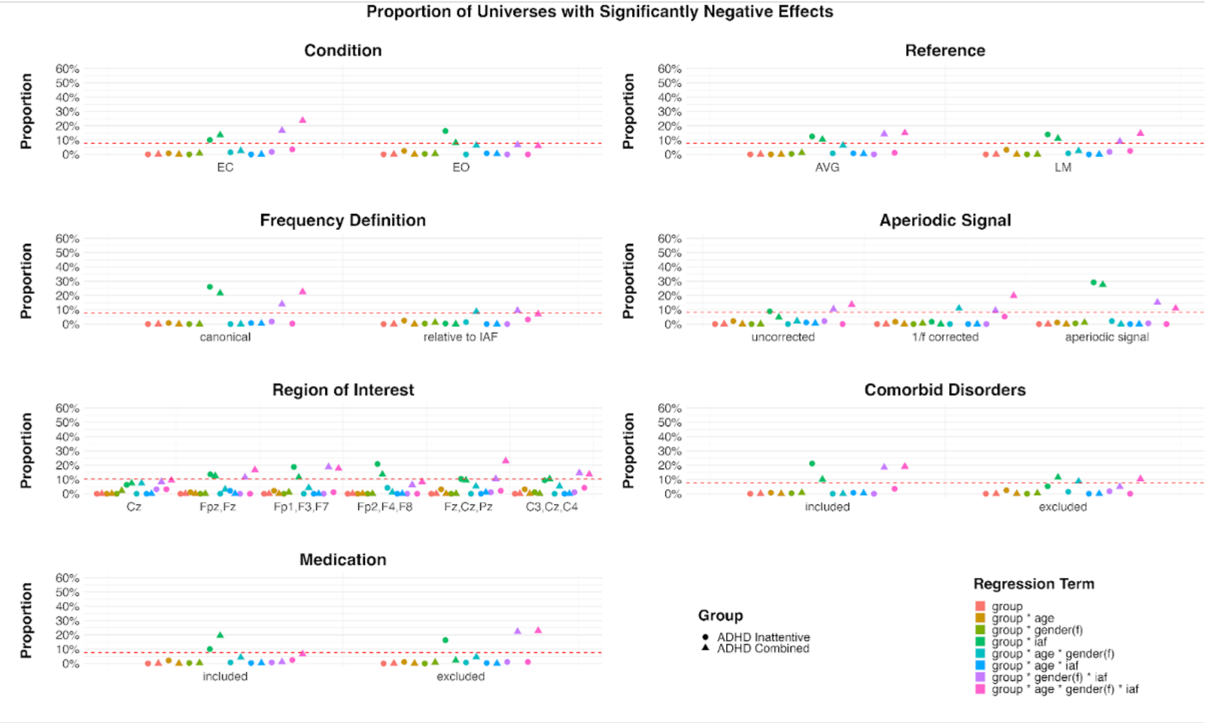


Supplementary Figure 3. Proportion of specifications showing a significantly negative effect by regression term and ADHD subtype across all analytical specifications. Each level of Condition, Reference, Frequency Definition, Comorbid Disorder, and Medication includes 288 analytical universes. The Aperiodic Signal factor includes 192 universes per level, and the Region of Interest includes 96 universes per level. The red line in the plot indicates the exact proportion of significant results required for the binomial test to reach statistical significance ($p < .05$) for each subset size.

##### 1.1.3. Supplementary Tables with Selected Models

To illustrate results from individual analysis universes in the HBN sample, we present six representative models. In all cases, TBR was computed from EO recordings at Cz with an average reference, including participants with comorbidities and medication. The models vary only in the choice of frequency band definition and spectral measure, thereby highlighting how these analytic decisions shape the observed effects.

Supplementary Table 1. Effects of Group, Age, Gender, and IAF on TBR calculated from 1/f-uncorrected power using canonical frequency bands.

| Variable | β | SE | CI | t-value | p-value |
|---|---|---|---|---|---|
| Intercept | 0.00 | 0.10 | -0.2 – 0.2 | 0.00 | p = 1.000 |
| Age | -0.10 | 0.09 | -0.29 – 0.08 | -1.09 | p = 0.277 |
| Gender[Female] | -0.09 | 0.15 | -0.38 – 0.2 | -0.59 | p = 0.557 |
| IAF | -0.13 | 0.11 | -0.36 – 0.09 | -1.16 | p = 0.246 |
| Age * Gender[Female] | 0.00 | 0.14 | -0.27 – 0.27 | 0.00 | p = 1.000 |
| Age * IAF | 0.00 | 0.10 | -0.2 – 0.2 | 0.02 | p = 0.986 |
| Gender[Female] * IAF | 0.08 | 0.16 | -0.23 – 0.39 | 0.51 | p = 0.611 |
| Age * Gender[Female] * IAF | -0.01 | 0.13 | -0.28 – 0.25 | -0.11 | p = 0.912 |
| ADHD[IN] | 0.00 | 0.12 | -0.23 – 0.23 | -0.01 | p = 0.995 |
| ADHD[IN] * Age | -0.07 | 0.11 | -0.29 – 0.16 | -0.58 | p = 0.563 |
| ADHD[IN] * Gender[Female] | -0.12 | 0.18 | -0.47 – 0.24 | -0.65 | p = 0.516 |
| ADHD[IN] * IAF | -0.02 | 0.13 | -0.27 – 0.24 | -0.13 | p = 0.898 |
| ADHD[IN] * Age * Gender[Female] | 0.01 | 0.18 | -0.34 – 0.36 | 0.04 | p = 0.965 |
| ADHD[IN] * Age * IAF | 0.09 | 0.12 | -0.15 – 0.32 | 0.72 | p = 0.474 |
| ADHD[IN] * Gender[Female] * IAF | -0.01 | 0.19 | -0.38 – 0.37 | -0.03 | p = 0.972 |
| ADHD[IN] * Age * Gender[Female] * IAF | 0.01 | 0.18 | -0.35 – 0.36 | 0.04 | p = 0.964 |
| ADHD[Comb] | 0.14 | 0.12 | -0.09 – 0.37 | 1.23 | p = 0.220 |
| ADHD[Comb] * Age | 0.13 | 0.11 | -0.09 – 0.36 | 1.18 | p = 0.238 |
| ADHD[Comb] * Gender[Female] | -0.11 | 0.20 | -0.49 – 0.28 | -0.54 | p = 0.588 |
| ADHD[Comb] * IAF | -0.01 | 0.13 | -0.26 – 0.25 | -0.05 | p = 0.959 |
| ADHD[Comb] * Age * Gender[Female] | -0.13 | 0.19 | -0.51 – 0.25 | -0.67 | p = 0.502 |
| ADHD[Comb] * Age * IAF | -0.01 | 0.12 | -0.25 – 0.23 | -0.09 | p = 0.931 |
| ADHD[Comb] * Gender[Female] * IAF | -0.06 | 0.19 | -0.44 – 0.31 | -0.33 | p = 0.741 |
| ADHD[Comb] * Age * Gender[Female] * IAF | 0.06 | 0.17 | -0.28 – 0.4 | 0.37 | p = 0.709 |

Note. In = Inattentive. Comb = Combined. IAF = Individual Alpha Frequency. SE = Standard Error. CI = 95% Confidence Intervals.
**p < 0.05. **p < 0.01. ***p < 0.001*

Supplementary Table 2. Effects of Group, Age, Gender, and IAF on TBR calculated from 1/f-uncorrected power using frequency bands relative to IAF.

| Variable | β | SE | CI | t-value | p-value |
|---|---|---|---|---|---|
| Intercept | -0.09 | 0.07 | -0.22 – 0.05 | -1.22 | p = 0.222 |
| Age | -0.24 | 0.07 | -0.37 – -0.11 | -3.59 | p = 3.43e-04*** |
| Gender[Female] | -0.05 | 0.10 | -0.25 – 0.15 | -0.49 | p = 0.623 |
| IAF | -0.53 | 0.08 | -0.69 – -0.37 | -6.66 | p = 4.37e-11*** |
| Age * Gender[Female] | 0.06 | 0.10 | -0.13 – 0.24 | 0.58 | p = 0.559 |
| Age * IAF | 0.29 | 0.07 | 0.15 – 0.43 | 4.09 | p = 4.69e-05*** |
| Gender[Female] * IAF | -0.03 | 0.11 | -0.25 – 0.18 | -0.32 | p = 0.751 |
| Age * Gender[Female] * IAF | -0.14 | 0.09 | -0.32 – 0.04 | -1.50 | p = 0.135 |
| ADHD[IN] | 0.04 | 0.08 | -0.12 – 0.21 | 0.54 | p = 0.591 |
| ADHD[IN] * Age | 0.02 | 0.08 | -0.13 – 0.18 | 0.26 | p = 0.793 |
| ADHD[IN] * Gender[Female] | -0.04 | 0.13 | -0.28 – 0.21 | -0.28 | p = 0.779 |
| ADHD[IN] * IAF | -0.10 | 0.09 | -0.28 – 0.08 | -1.14 | p = 0.256 |
| ADHD[IN] * Age * Gender[Female] | 0.01 | 0.13 | -0.24 – 0.25 | 0.04 | p = 0.965 |
| ADHD[IN] * Age * IAF | -0.11 | 0.08 | -0.27 – 0.05 | -1.31 | p = 0.189 |
| ADHD[IN] * Gender[Female] * IAF | 0.07 | 0.13 | -0.2 – 0.33 | 0.49 | p = 0.624 |
| ADHD[IN] * Age * Gender[Female] * IAF | 0.16 | 0.13 | -0.09 – 0.41 | 1.25 | p = 0.213 |
| ADHD[Comb] | 0.08 | 0.08 | -0.08 – 0.24 | 1.00 | p = 0.316 |
| ADHD[Comb] * Age | 0.17 | 0.08 | 0.02 – 0.33 | 2.18 | p = 0.030* |
| ADHD[Comb] * Gender[Female] | 0.02 | 0.14 | -0.25 – 0.29 | 0.18 | p = 0.860 |
| ADHD[Comb] * IAF | -0.12 | 0.09 | -0.3 – 0.06 | -1.35 | p = 0.177 |
| ADHD[Comb] * Age * Gender[Female] | -0.05 | 0.14 | -0.32 – 0.21 | -0.39 | p = 0.696 |
| ADHD[Comb] * Age * IAF | -0.17 | 0.08 | -0.34 – -0.01 | -2.06 | p = 0.040* |
| ADHD[Comb] * Gender[Female] * IAF | 0.02 | 0.14 | -0.25 – 0.28 | 0.14 | p = 0.888 |
| ADHD[Comb] * Age * Gender[Female] * IAF | 0.15 | 0.12 | -0.09 – 0.39 | 1.26 | p = 0.209 |

Note. In = Inattentive. Comb = Combined. IAF = Individual Alpha Frequency. SE = Standard Error. CI = 95% Confidence Intervals.

**p < 0.05. **p < 0.01. ***p < 0.001*

Supplementary Table 3. Effects of Group, Age, Gender, and IAF on TBR calculated from aperiodic-adjusted power using canonical frequency bands.

| Variable | β | SE | CI | t-value | p-value |
|---|---|---|---|---|---|
| Intercept | -0.01 | 0.10 | -0.21 – 0.18 | -0.15 | p = 0.882 |
| Age | -0.05 | 0.10 | -0.23 – 0.14 | -0.50 | p = 0.618 |
| Gender[Female] | -0.08 | 0.15 | -0.37 – 0.21 | -0.54 | p = 0.591 |
| IAF | -0.12 | 0.11 | -0.34 – 0.1 | -1.05 | p = 0.292 |
| Age * Gender[Female] | -0.02 | 0.14 | -0.28 – 0.25 | -0.13 | p = 0.899 |
| Age * IAF | -0.06 | 0.10 | -0.26 – 0.14 | -0.58 | p = 0.559 |
| Gender[Female] * IAF | 0.03 | 0.16 | -0.27 – 0.34 | 0.22 | p = 0.825 |
| Age * Gender[Female] * IAF | 0.10 | 0.13 | -0.16 – 0.36 | 0.73 | p = 0.466 |
| ADHD[IN] | 0.00 | 0.12 | -0.23 – 0.23 | 0.01 | p = 0.994 |
| ADHD[IN] * Age | -0.06 | 0.11 | -0.28 – 0.16 | -0.55 | p = 0.585 |
| ADHD[IN] * Gender[Female] | -0.05 | 0.18 | -0.41 – 0.3 | -0.28 | p = 0.780 |
| ADHD[IN] * IAF | -0.09 | 0.13 | -0.35 – 0.16 | -0.73 | p = 0.467 |
| ADHD[IN] * Age * Gender[Female] | 0.07 | 0.18 | -0.28 – 0.42 | 0.40 | p = 0.691 |
| ADHD[IN] * Age * IAF | 0.15 | 0.12 | -0.08 – 0.38 | 1.26 | p = 0.208 |
| ADHD[IN] * Gender[Female] * IAF | 0.11 | 0.19 | -0.26 – 0.49 | 0.59 | p = 0.552 |
| ADHD[IN] * Age * Gender[Female] * IAF | -0.15 | 0.18 | -0.51 – 0.2 | -0.84 | p = 0.402 |
| ADHD[Comb] | 0.10 | 0.12 | -0.13 – 0.33 | 0.84 | p = 0.400 |
| ADHD[Comb] * Age | 0.08 | 0.11 | -0.15 – 0.3 | 0.67 | p = 0.503 |
| ADHD[Comb] * Gender[Female] | 0.03 | 0.20 | -0.35 – 0.42 | 0.18 | p = 0.859 |
| ADHD[Comb] * IAF | -0.02 | 0.13 | -0.28 – 0.23 | -0.16 | p = 0.873 |
| ADHD[Comb] * Age * Gender[Female] | -0.06 | 0.19 | -0.44 – 0.32 | -0.32 | p = 0.751 |
| ADHD[Comb] * Age * IAF | 0.09 | 0.12 | -0.14 – 0.33 | 0.78 | p = 0.434 |
| ADHD[Comb] * Gender[Female] * IAF | -0.15 | 0.19 | -0.52 – 0.23 | -0.75 | p = 0.452 |
| ADHD[Comb] * Age * Gender[Female] * IAF | -0.11 | 0.17 | -0.45 – 0.24 | -0.61 | p = 0.544 |

Note. In = Inattentive. Comb = Combined. IAF = Individual Alpha Frequency. SE = Standard Error. CI = 95% Confidence Intervals.
**p < 0.05. **p < 0.01. ***p < 0.001*

Supplementary Table 4. Effects of Group, Age, Gender, and IAF on TBR calculated from aperiodic-adjusted power using frequency bands relative to IAF.

| Variable | β | SE | CI | t-value | p-value |
|---|---|---|---|---|---|
| Intercept | -0.06 | 0.10 | -0.26 – 0.13 | -0.63 | p = 0.528 |
| Age | -0.23 | 0.09 | -0.42 – -0.04 | -2.43 | p = 0.015* |
| Gender[Female] | 0.09 | 0.15 | -0.19 – 0.38 | 0.64 | p = 0.522 |
| IAF | 0.25 | 0.11 | 0.03 – 0.47 | 2.18 | p = 0.029* |
| Age * Gender[Female] | 0.08 | 0.14 | -0.19 – 0.34 | 0.55 | p = 0.580 |
| Age * IAF | -0.03 | 0.10 | -0.23 – 0.16 | -0.34 | p = 0.736 |
| Gender[Female] * IAF | -0.23 | 0.16 | -0.54 – 0.08 | -1.47 | p = 0.142 |
| Age * Gender[Female] * IAF | 0.09 | 0.13 | -0.17 – 0.35 | 0.66 | p = 0.508 |
| ADHD[IN] | 0.03 | 0.12 | -0.2 – 0.26 | 0.26 | p = 0.793 |
| ADHD[IN] * Age | 0.01 | 0.11 | -0.22 – 0.23 | 0.05 | p = 0.960 |
| ADHD[IN] * Gender[Female] | 0.05 | 0.18 | -0.3 – 0.41 | 0.29 | p = 0.773 |
| ADHD[IN] * IAF | -0.09 | 0.13 | -0.34 – 0.17 | -0.67 | p = 0.502 |
| ADHD[IN] * Age * Gender[Female] | 0.07 | 0.18 | -0.27 – 0.42 | 0.42 | p = 0.678 |
| ADHD[IN] * Age * IAF | 0.02 | 0.12 | -0.22 – 0.25 | 0.13 | p = 0.896 |
| ADHD[IN] * Gender[Female] * IAF | 0.05 | 0.19 | -0.32 – 0.42 | 0.26 | p = 0.797 |
| ADHD[IN] * Age * Gender[Female] * IAF | -0.04 | 0.18 | -0.39 – 0.32 | -0.21 | p = 0.837 |
| ADHD[Comb] | 0.12 | 0.12 | -0.11 – 0.35 | 1.00 | p = 0.319 |
| ADHD[Comb] * Age | 0.21 | 0.11 | -0.02 – 0.43 | 1.82 | p = 0.069 |
| ADHD[Comb] * Gender[Female] | -0.12 | 0.20 | -0.5 – 0.27 | -0.60 | p = 0.548 |
| ADHD[Comb] * IAF | 0.04 | 0.13 | -0.22 – 0.29 | 0.29 | p = 0.774 |
| ADHD[Comb] * Age * Gender[Female] | -0.15 | 0.19 | -0.52 – 0.23 | -0.76 | p = 0.449 |
| ADHD[Comb] * Age * IAF | 0.03 | 0.12 | -0.2 – 0.27 | 0.27 | p = 0.785 |
| ADHD[Comb] * Gender[Female] * IAF | 0.05 | 0.19 | -0.32 – 0.43 | 0.29 | p = 0.775 |
| ADHD[Comb] * Age * Gender[Female] * IAF | -0.13 | 0.17 | -0.47 – 0.21 | -0.75 | p = 0.451 |

Note. In = Inattentive. Comb = Combined. IAF = Individual Alpha Frequency. SE = Standard Error. CI = 95% Confidence Intervals.
**p < 0.05. **p < 0.01. ***p < 0.001*

Supplementary Table 5. Effects of Group, Age, Gender, and IAF on TBR calculated from aperiodic signal power using canonical frequency bands.

| Variable | β | SE | CI | t-value | p-value |
|---|---|---|---|---|---|
| Intercept | 0.13 | 0.09 | -0.05 – 0.32 | 1.40 | p = 0.162 |
| Age | -0.20 | 0.09 | -0.37 – -0.02 | -2.20 | p = 0.028* |
| Gender[Female] | -0.30 | 0.14 | -0.57 – -0.03 | -2.15 | p = 0.032* |
| IAF | -0.27 | 0.11 | -0.48 – -0.06 | -2.51 | p = 0.012* |
| Age * Gender[Female] | 0.01 | 0.13 | -0.24 – 0.26 | 0.06 | p = 0.953 |
| Age * IAF | 0.10 | 0.10 | -0.09 – 0.28 | 1.02 | p = 0.309 |
| Gender[Female] * IAF | 0.24 | 0.15 | -0.04 – 0.53 | 1.66 | p = 0.098 |
| Age * Gender[Female] * IAF | -0.25 | 0.12 | -0.49 – -0.01 | -2.01 | p = 0.045* |
| ADHD[IN] | -0.01 | 0.11 | -0.23 – 0.2 | -0.11 | p = 0.911 |
| ADHD[IN] * Age | -0.08 | 0.11 | -0.29 – 0.13 | -0.75 | p = 0.456 |
| ADHD[IN] * Gender[Female] | -0.33 | 0.17 | -0.66 – 0 | -1.95 | p = 0.052 |
| ADHD[IN] * IAF | 0.16 | 0.12 | -0.08 – 0.4 | 1.33 | p = 0.185 |
| ADHD[IN] * Age * Gender[Female] | -0.08 | 0.17 | -0.41 – 0.25 | -0.49 | p = 0.622 |
| ADHD[IN] * Age * IAF | -0.03 | 0.11 | -0.25 – 0.18 | -0.31 | p = 0.757 |
| ADHD[IN] * Gender[Female] * IAF | -0.22 | 0.18 | -0.57 – 0.13 | -1.23 | p = 0.220 |
| ADHD[IN] * Age * Gender[Female] * IAF | 0.33 | 0.17 | 0 – 0.66 | 1.93 | p = 0.053 |
| ADHD[Comb] | 0.10 | 0.11 | -0.12 – 0.31 | 0.90 | p = 0.367 |
| ADHD[Comb] * Age | 0.02 | 0.11 | -0.19 – 0.23 | 0.15 | p = 0.882 |
| ADHD[Comb] * Gender[Female] | -0.27 | 0.18 | -0.63 – 0.09 | -1.49 | p = 0.136 |
| ADHD[Comb] * IAF | 0.13 | 0.12 | -0.11 – 0.37 | 1.07 | p = 0.283 |
| ADHD[Comb] * Age * Gender[Female] | -0.03 | 0.18 | -0.38 – 0.33 | -0.14 | p = 0.886 |
| ADHD[Comb] * Age * IAF | -0.14 | 0.11 | -0.36 – 0.08 | -1.27 | p = 0.205 |
| ADHD[Comb] * Gender[Female] * IAF | -0.13 | 0.18 | -0.48 – 0.23 | -0.71 | p = 0.475 |
| ADHD[Comb] * Age * Gender[Female] * IAF | 0.43 | 0.16 | 0.11 – 0.75 | 2.66 | p = 0.008** |

Note. In = Inattentive. Comb = Combined. IAF = Individual Alpha Frequency. SE = Standard Error. CI = 95% Confidence Intervals.
**p < 0.05. **p < 0.01. ***p < 0.001*

Supplementary Table 6. Effects of Group, Age, Gender, and IAF on TBR calculated from aperiodic signal power using frequency bands relative to IAF.

| Variable | β | SE | CI | t-value | p-value |
|---|---|---|---|---|---|
| Intercept | -0.14 | 0.06 | -0.25 – -0.03 | -2.54 | p = 0.011* |
| Age | -0.14 | 0.05 | -0.24 – -0.04 | -2.65 | p = 0.008** |
| Gender[Female] | -0.03 | 0.08 | -0.19 – 0.13 | -0.34 | p = 0.737 |
| IAF | -0.69 | 0.06 | -0.81 – -0.56 | -10.80 | p = 6.23e-26*** |
| Age * Gender[Female] | 0.04 | 0.08 | -0.11 – 0.19 | 0.56 | p = 0.576 |
| Age * IAF | 0.33 | 0.06 | 0.22 – 0.44 | 5.80 | p = 8.66e-09*** |
| Gender[Female] * IAF | 0.11 | 0.09 | -0.06 – 0.28 | 1.27 | p = 0.205 |
| Age * Gender[Female] * IAF | -0.17 | 0.07 | -0.32 – -0.02 | -2.29 | p = 0.022* |
| ADHD[IN] | 0.08 | 0.07 | -0.05 – 0.21 | 1.23 | p = 0.220 |
| ADHD[IN] * Age | 0.04 | 0.06 | -0.08 – 0.17 | 0.67 | p = 0.505 |
| ADHD[IN] * Gender[Female] | -0.06 | 0.10 | -0.26 – 0.14 | -0.57 | p = 0.567 |
| ADHD[IN] * IAF | -0.11 | 0.07 | -0.25 – 0.03 | -1.55 | p = 0.121 |
| ADHD[IN] * Age * Gender[Female] | -0.02 | 0.10 | -0.22 – 0.18 | -0.20 | p = 0.841 |
| ADHD[IN] * Age * IAF | -0.09 | 0.07 | -0.22 – 0.04 | -1.31 | p = 0.190 |
| ADHD[IN] * Gender[Female] * IAF | 0.08 | 0.11 | -0.13 – 0.29 | 0.76 | p = 0.446 |
| ADHD[IN] * Age * Gender[Female] * IAF | 0.06 | 0.10 | -0.14 – 0.26 | 0.61 | p = 0.545 |
| ADHD[Comb] | 0.10 | 0.07 | -0.03 – 0.23 | 1.50 | p = 0.134 |
| ADHD[Comb] * Age | 0.10 | 0.06 | -0.03 – 0.22 | 1.49 | p = 0.136 |
| ADHD[Comb] * Gender[Female] | 0.14 | 0.11 | -0.08 – 0.35 | 1.25 | p = 0.213 |
| ADHD[Comb] * IAF | -0.16 | 0.07 | -0.3 – -0.01 | -2.14 | p = 0.033* |
| ADHD[Comb] * Age * Gender[Female] | 0.07 | 0.11 | -0.15 – 0.28 | 0.61 | p = 0.540 |
| ADHD[Comb] * Age * IAF | -0.13 | 0.07 | -0.26 – 0 | -1.96 | p = 0.051 |
| ADHD[Comb] * Gender[Female] * IAF | -0.11 | 0.11 | -0.32 – 0.1 | -1.00 | p = 0.317 |
| ADHD[Comb] * Age * Gender[Female] * IAF | 0.06 | 0.10 | -0.13 – 0.25 | 0.62 | p = 0.536 |

Note. In = Inattentive. Comb = Combined. IAF = Individual Alpha Frequency. SE = Standard Error. CI = 95% Confidence Intervals.
**p < 0.05. **p < 0.01. ***p < 0.001*

##### 1.1.4. Bootstrap Robustness Analysis

To evaluate the robustness of our multiverse findings to sample size imbalances, we conducted a bootstrap-based sensitivity analysis. For each universe, we identified the group with the lowest number of participants (e.g., ADHD-Inattentive, female, no medication, no comorbidities) and randomly resampled all groups to this minimum size. The multiverse analysis was then re-run on these balanced subsamples. This procedure was repeated 1000 times with independent random draws. The bootstrap analysis yielded results that closely mirrored the main multiverse findings. The frequency and direction of significant effects were consistent across resampled iterations, indicating that our conclusions were not driven by unequal group sizes or by idiosyncratic subsamples (Supplementary Table 8).

Supplementary Table 8. Results of the bootstrap robustness analysis across 1000 resampled multiverse iterations.

| | ***Significantly Positive*** | ***Significantly Negative*** | ***Not Significant*** | ***Total*** |
|---|---|---|---|---|
| ADHD[IN] | 13.28 ± 3.5 (2.31%) | 15.86 ± 3.58 (2.75 %) | 546.86 ± 5.13 (94.94 %) | 576 |
| ADHD[IN] * Age | 10.17 ± 3.15 (1.77 %) | 17.15 ± 3.77 (2.98 %) | 548.68 ± 4.92 (95.26 %) | 576 |
| ADHD[IN] * Gender[Female] | 20.68 ± 3.33 (3.59 %) | 6.68 ± 2.43 (1.16 %) | 548.64 ± 4.17 (95.25 %) | 576 |
| ADHD[IN] * IAF | 23.90 ± 4.75 (4.15 %) | 61.40 ± 6.42 (10.66 %) | 490.70 ± 7.78 (85.19 %) | 576 |
| ADHD[IN] * Age * Gender[Female] | 4.20 ± 1.99 (0.73 %) | 6.16 ± 2.25 (1.07 %) | 565.64 ± 2.96 (98.20 %) | 576 |
| ADHD[IN] * Age * IAF | 62.80 ± 6.63 (10.90 %) | 30.16 ± 5.03 (5.24 %) | 483.04 ± 8.01 (83.86 %) | 576 |
| ADHD[IN] * Gender[Female] * IAF | 40.34 ± 4.75 (7.00 %) | 18.59 ± 4.03 (3.23 %) | 517.07 ± 6.12 (89.77 %) | 576 |
| ADHD[IN] * Age * Gender[Female] * IAF | 22.90 ± 4.47 (3.98 %) | 27.34 ± 4.67 (4.75 %) | 525.76 ± 6.46 (91.28 %) | 576 |
| ADHD[Comb] | 28.03 ± 4.76 (4.87 %) | 4.06 ± 2.00 (0.70 %) | 543.92 ± 5.13 (94.43 %) | 576 |
| ADHD[Comb] * Age | 51.01 ± 6.32 (8.86 %) | 6.30 ± 2.44 (1.09 %) | 518.70 ± 6.80 (90.05 %) | 576 |
| ADHD[Comb] * Gender[Female] | 56.06 ± 5.82 (9.73 %) | 4.25 ± 1.97 (0.74 %) | 515.69 ± 6.10 (89.53 %) | 576 |

| | | | | |
|---|---|---|---|---|
| ADHD[Comb] * IAF | 12.01 ± 3.42 (2.09 %) | 46.67 ± 6.07 (8.10 %) | 517.32 ± 6.87 (89.81 %) | 576 |
| ADHD[Comb] * Age * Gender[Female] | 44.40 ± 4.94 (7.71 %) | 15.09 ± 3.30 (2.62 %) | 516.51 ± 5.95 (89.67 %) | 576 |
| ADHD[Comb] * Age * IAF | 44.38 ± 5.76 (7.70 %) | 27.80 ± 4.87 (4.83 %) | 503.83 ± 7.10 (87.47 %) | 576 |
| ADHD[Comb] * Gender[Female] * IAF | 12.76 ± 3.25 (2.22 %) | 64.78 ± 5.87 (11.25 %) | 498.46 ± 6.68 (86.54 %) | 576 |
| ADHD[Comb] * Age * Gender[Female] * IAF | 9.99 ± 2.80 (1.74 %) | 87.31 ± 7.41 (15.16 %) | 478.70 ± 7.85 (83.11 %) | 576 |

Note. Values reported represent the mean across 1000 iterations ± standard deviation, as well as the percentage reflecting the mean value. Positive universe = higher TBR in ADHD compared to HC. Negative universe = lower TBR in ADHD compared to HC. IN = Inattentive. Comb = Combined. IAF = individual alpha frequency.

##### 1.1.5. Dimensional multiverse analysis with SWAN Total scores

To complement the categorical analyses based on the diagnostic group (i.e., HC, ADHD-Inattentive, ADHD-Combined), we conducted an identical multiverse analysis using the SWAN scores [1] as a dimensional predictor of ADHD symptom severity in the HBN sample only, as SWAN scores were not available for the validation sample. This dimensional approach tested the robustness of findings across the full spectrum of ADHD symptomatology, avoiding potential information loss from diagnostic thresholds while increasing statistical power and capturing subclinical variability. The identical set of analytical specifications and model structures were applied, substituting the categorical Group factor with continuous SWAN scores and their interactions with age, gender, and IAF.

Supplementary Table 7. Results of the multiverse analysis using dimensional SWAN scores instead of categorical ADHD diagnoses.

| | ***Significantly Positive*** | ***Significantly Negative*** | ***Not Significant*** | ***Total*** |
|---|---|---|---|---|
| SWAN | 0 (0.00 %) | 21 (3.65 %) | 555 (96.35 %) | 576 |
| SWAN * Age | 7 (1.21 %) | 20 (3.47 %) | 549 (95.31 %) | 576 |
| SWAN * Gender[Female] | 65 (11.28%) | 0 (0.00%) | 562 (88.72 %) | 576 |
| SWAN * IAF | 14 (2.43 %) | 134 (23.26 %) | 428 (74.31 %) | 576 |
| SWAN * Age * Gender[Female] | 9 (1.56 %) | 0 (0.00 %) | 567 (98.44 %) | 576 |
| SWAN * Age * IAF | 43 (7.47%) | 0 (0.00 %) | 533 (92.53 %) | 576 |
| SWAN * Gender[Female] * IAF | 79 (13.71 %) | 5 (0.87 %) | 492 (85.42 %) | 576 |
| SWAN * Age * Gender[Female] * IAF | 3 (0.52 %) | 67 (11.63 %) | 506 (87.85 %) | 576 |

Note. Positive universe = higher TBR in ADHD compared to HC. Negative universe = lower TBR in ADHD compared to HC. IAF = individual alpha frequency.

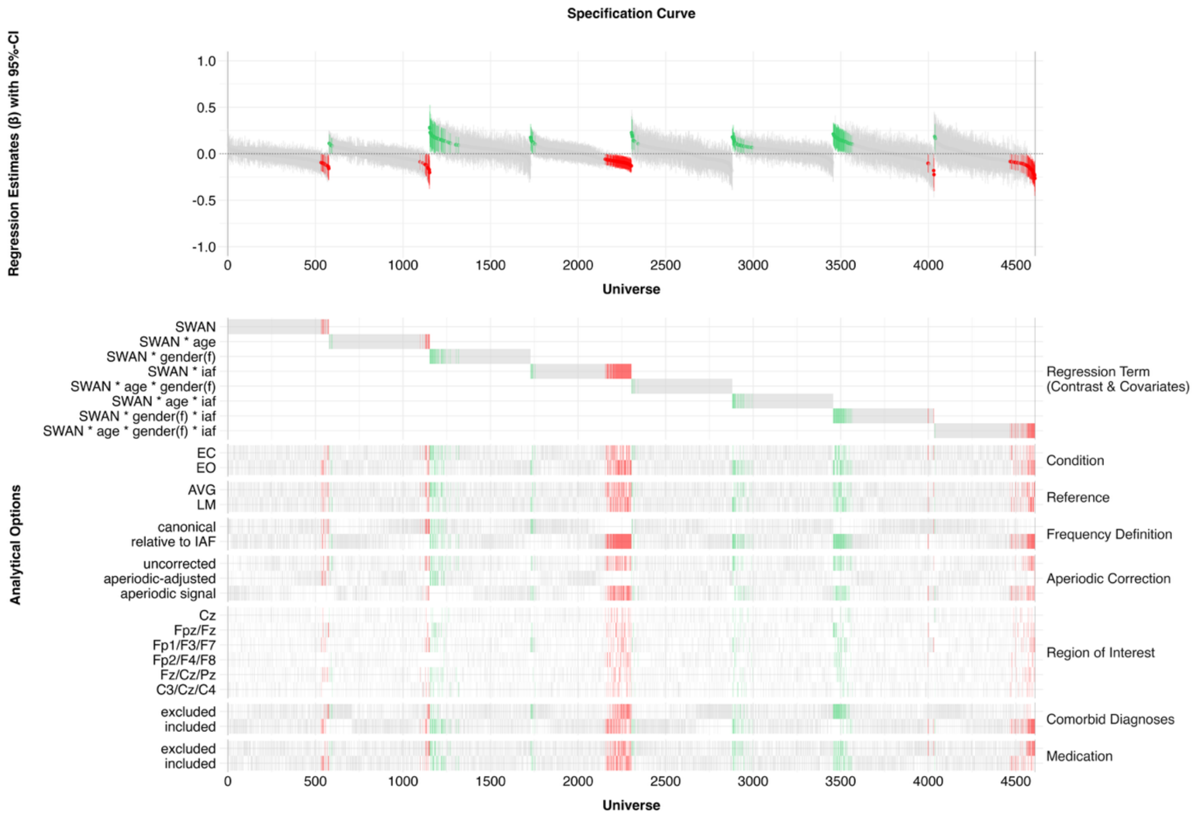


Supplementary Figure 3. Specification curve representing all universes for the dimensional SWAN score analysis. The top panel shows regression estimates for each specification, sorted by regression estimate, with 95% confidence intervals. Statistically significant positive estimates are shown in green, negative estimates in red, and non-significant estimates in gray. The bottom panel indicates which analytical choices were associated with significantly positive (i.e., higher TBR for ADHD compared to HC) or negative (i.e., lower TBR for ADHD compared to HC) effects across specifications. Note. CI = confidence interval. f = female. EC = eyes closed. EO = eyes open. AVG = average reference. LM = linked mastoid reference. Canonical = canonical frequency range. Relative to IAF = bandwidth relative to individual alpha frequency.

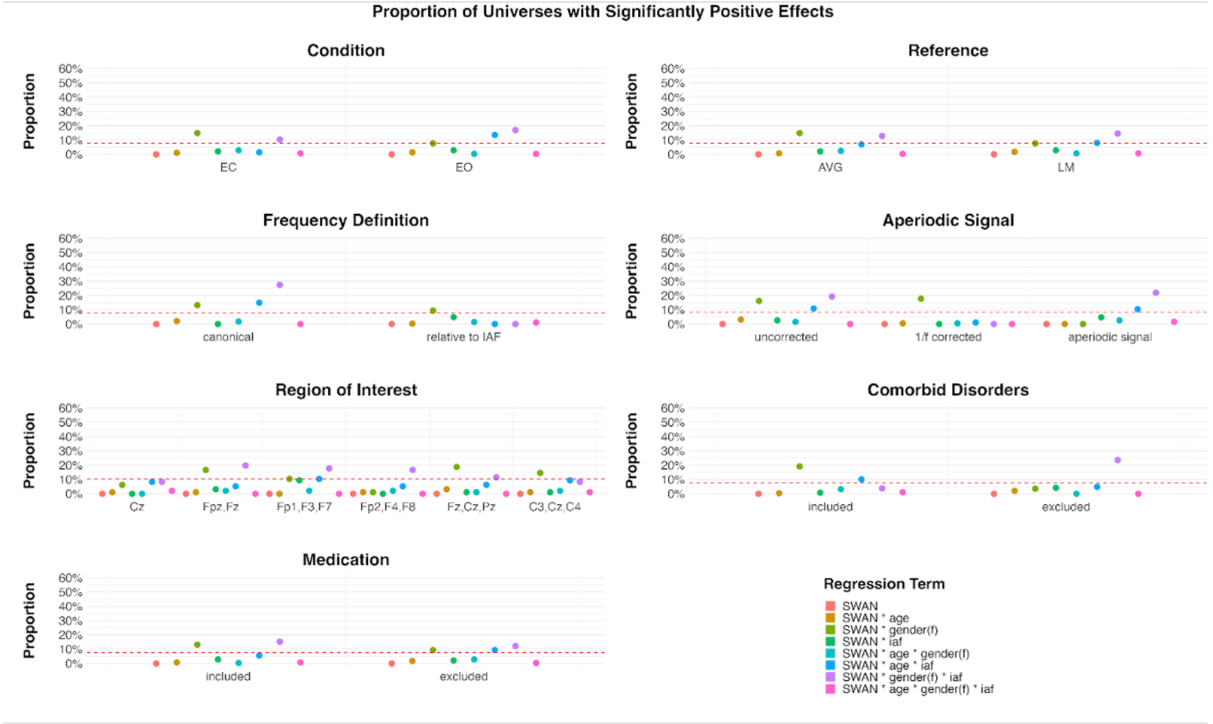

Supplementary Figure 4. Proportion of specifications showing a significantly positive effect by regression term across all analytical specifications. Each level of Condition, Reference, Frequency Definition, Comorbid Disorder, and Medication includes 288 analytical universes. The Aperiodic Signal factor includes 192 universes per level, and the Region of Interest includes 96 universes per level. The red line in the plot indicates the exact proportion of significant results required for the binomial test to reach statistical significance ($p < .05$) for each subset size.

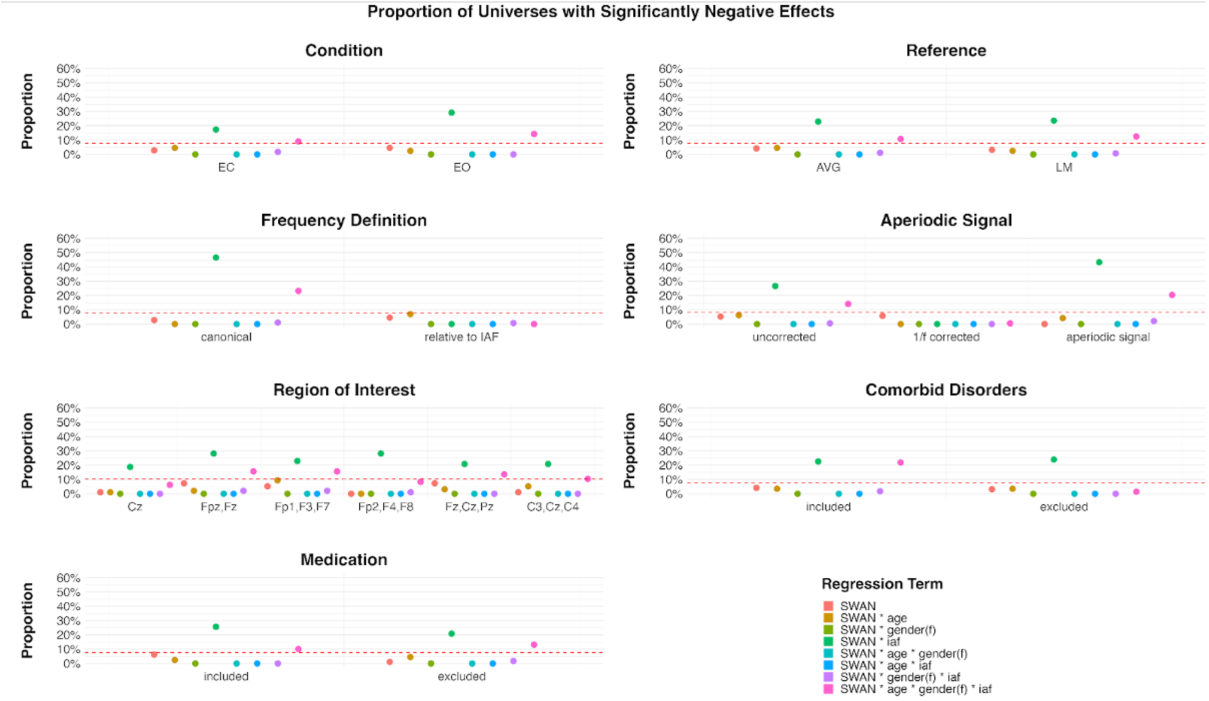


Supplementary Figure 5. Proportion of specifications showing a significantly negative effect by regression term across all analytical specifications. Each level of Condition, Reference, Frequency Definition, Comorbid Disorder, and Medication includes 288 analytical universes. The Aperiodic Signal factor includes 192 universes per level, and the Region of Interest includes 96 universes per level. The red line in the plot indicates the exact proportion of significant results required for the binomial test to reach statistical significance ($p < .05$) for each subset size.

#### 1.2. Validation Sample
##### 1.2.1. Methods
###### 1.2.1.1. Participants

The second dataset used for validation was obtained from a previously conducted multicenter study carried out as part of the project coordinated by the Brain and Trauma Foundation Grisons, Switzerland [2-4]. The dataset comprised 381 participants, including both individuals with ADHD and healthy controls. The sample consisted of 191 healthy controls (mean age = 14.23 years, SD = 4.39, range = 6.28-21.90), 110 participants with ADHD-Inattentive Type (mean age = 12.88 years, SD = 3.35, range = 6.48-21.81), 65 with ADHD-Combined Type (mean age = 11.19 years, SD = 3.73, range = 6.28-21.35), and 15 with ADHD-Hyperactive/Impulsive Type (mean age = 10.34 years, SD = 3.55, range = 6.39-19.69). Due to the small sample size of the ADHD-Hyperactive/Impulsive group, it was excluded from further analyses.

Prior to participation, interested individuals completed an ADHD screening questionnaire. During the initial appointment, informed consent was obtained and a DSM-5-based ADHD diagnosis was made or confirmed by board-certified psychiatrists or clinical psychologists. The first testing session included structured clinical interviews (SKID; [5]), IQ testing using age-appropriate instruments (CFT 1-R or CFT 20-R: [6]; WMT-2: [7]), and a battery of neuropsychological tests (candit.com). EEG data were recorded in a subsequent session. General exclusion criteria included a history of traumatic brain injury, loss of consciousness, major psychiatric disorders (e.g., depression or schizophrenia), drug abuse, epilepsy, pregnancy, and an IQ below 80. All participants were required to be fluent in German, capable of understanding study procedures, and able to provide informed consent. While all participants were medication-free at the time of EEG recording, 139 ADHD participants were regularly treated with methylphenidate in daily life (e.g., Ritalin, Concerta, Elvanse). For detailed demographic information, see Supplementary Table 9. The study was approved by the cantonal ethics committee of Zurich (KEK-ZH 2013-0327 / EKNZ 2014-160), and data collection took place between July 2014 and July 2017 at five locations across Switzerland (Zürich, Chur, Lausanne, Lucerne, and Rapperswil).

Please note that detailed information on comorbid diagnoses and medication status was available only in the HBN sample; accordingly, these factors were examined exclusively in HBN. In the validation dataset, we knew that some participants were medicated but lacked details on compound, dose, or timing because this was a secondary dataset not collected by us and the information was not accessible. As a result, the specification set for the validation sample omitted the comorbidity and medication dimensions, reducing the number of analytic universes to 144 per group contrast (vs. 576 in HBN).

###### 1.2.1.2. EEG Acquisition

In the validation dataset, resting state EEG was recorded using a 19-channel system (NeuroAmp ×23) in combination with the ERPrec software (BEE Medic GmbH). EEG data were sampled at 500 Hz and recorded during a total of 8 minutes of resting-state recording, consisting of 4 minutes with EO and 4 minutes with EC. Electrodes were positioned according to the international 10–20 system using an electrode cap equipped with tin electrodes (Electro-Cap International Inc., Eaton, OH, USA). The recording reference was set to the linked earlobes, and electrode impedances were maintained below 5 kΩ. To ensure consistency with the main dataset (i.e. Healthy Brain Network), all EEG recordings were downsampled to 250 Hz prior to further processing.

##### 1.2.2. Results

###### 1.2.2.1. Demographics

Supplementary Table 9 and Figure 6 summarize the demographic characteristics of the validation sample after all exclusions were applied, as described in the Methods section. The final sample comprised 237 participants. Similar to the main sample, we first compared demographic characteristics of all three groups (i.e., HC, ADHD-Combined, ADHD-Inattentive). The chi-square test revealed a significant association between gender and diagnosis group, indicating that gender distribution differed across the three groups ($\chi^2 = 41.36$, $p$ = 1e-9). Next, the ANOVAs revealed significant group effects for age ($F = 11.35$, $p$ = 1.96e-5), and IQ ($F = 11.42$, $p$ = 1.87e-5), indicating that the HC, ADHD-Combined and ADHD-Inattentive differed significantly across these variables. The effect of the group for IAF was not statistically significant ($F = 2.47$, $p = 0.087$), suggesting that the groups did not differ in their IAF.

Supplementary Table 9. Demographics information of the validation sample.

| | *HC* | *Combined* | *Inattentive* | *$X^2$* | *p-value* |
|---|---|---|---|---|---|
| **N** | 107 | 46 | 84 | | |

| | | | | | | | | |
|---|---|---|---|---|---|---|---|---|
| Male | 45 | | 34 | | 72 | | | |
| Female | 62 | | 12 | | 12 | | 41.36 | 1e-9*** |
| | ***Mean*** | ***SD*** | ***Mean*** | ***SD*** | ***Mean*** | ***SD*** | ***F-value*** | ***p-value*** |
| **Age (years)** | 14.49 | 4.64 | 11.20 | 3.86 | 12.96 | 3.07 | 11.35 | 1.96e-5*** |
| **IQ** | 111.07 | 11.82 | 103.77 | 14.50 | 101.90 | 15.53 | 11.42 | 1.87e-5*** |
| **IAF** | 9.91 | 1.15 | 9.57 | 1.04 | 9.60 | 1.13 | 2.47 | 0.087 |

Note. HC = Healthy control. SD = Standard deviation. IN = Inattentive. HY = Hyperactive.
**p < 0.05. **p < 0.01. ***p < 0.001*

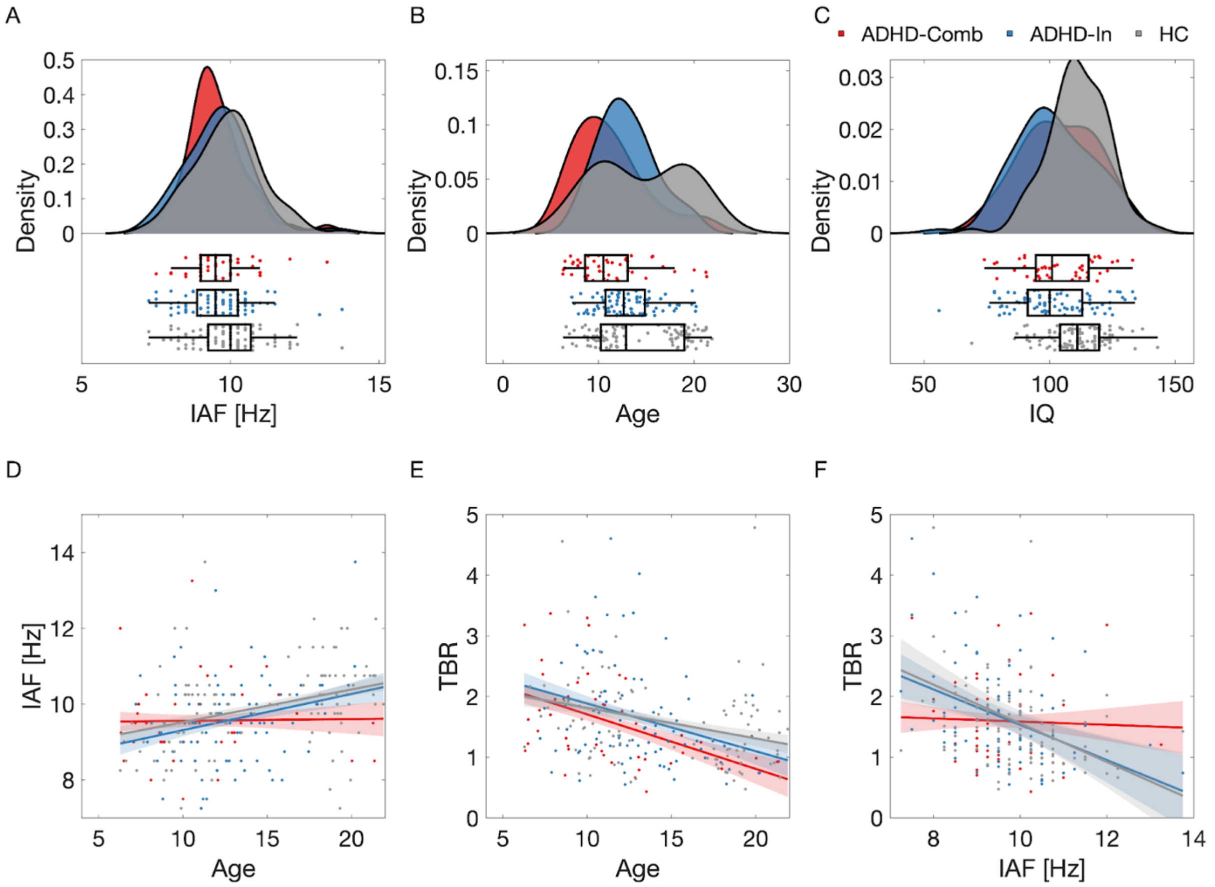


Supplementary Figure 6. Overview of sample characteristics and variable distributions. Density plots show the distribution of (A) IAF, (B) age and (C) IQ across the three groups. (D-F) Scatter plots display the relationship between (D) IAF and age, (E) TBR and age, (F) and TBR and IAF across all participants. The regression line is shown with standard error bands. Note. IAF = Individual Alpha Frequency. TBR = Theta-Beta Ratio. HC = Healthy Control.

#### 1.2.2.2. EEG features

##### 1.2.2.2.1. Aperiodic-adjusted power

To visualize the neurophysiological data, we plotted the scalp topographies of theta, beta and TBR as well as power spectra and aperiodic signal for HC, ADHD-Combined, and ADHD-

Inattentive groups (Supplementary Figure 7). The figure displays aperiodic-adjusted power during EO condition, computed using a canonical frequency range (theta: 4-8 Hz; beta: 13-30 Hz).

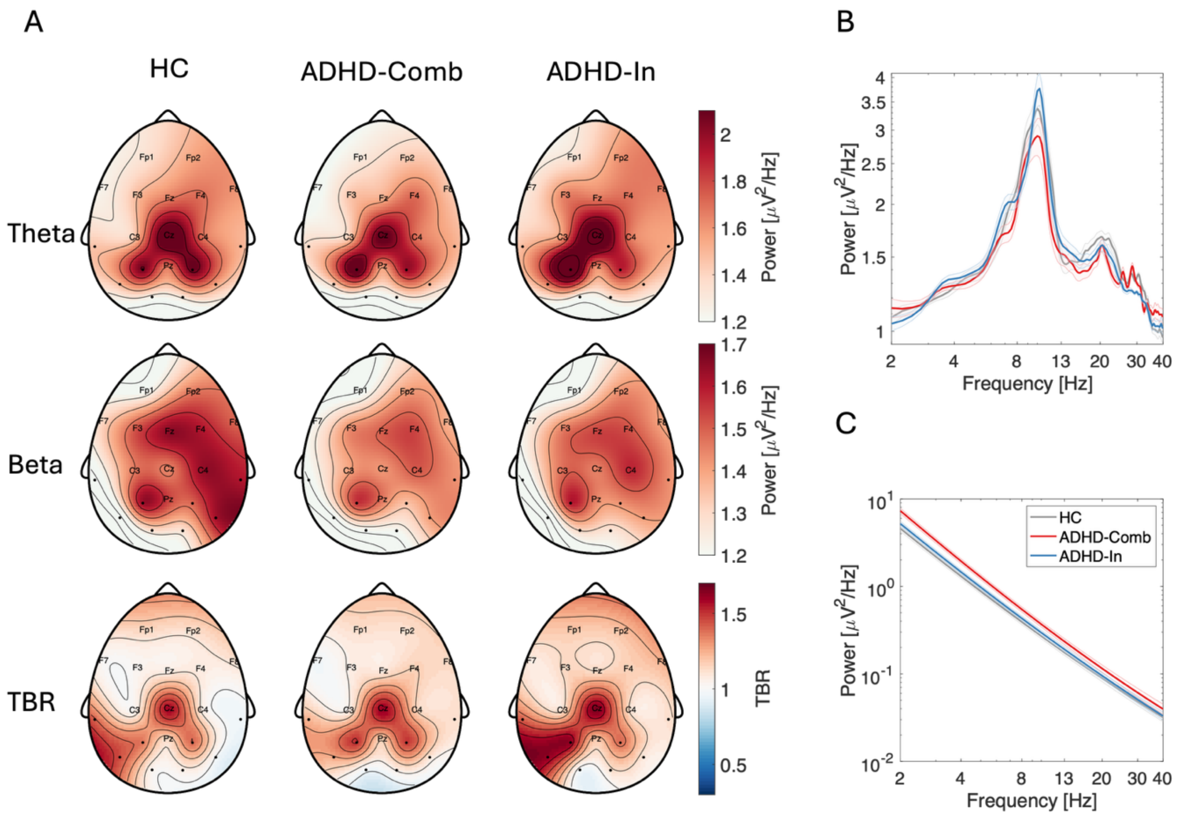


Supplementary Figure 7. Neurophysiological data from the validation sample. (A) Scalp topographies, (B) aperiodic-adjusted power spectra and (C) aperiodic signal for HC, ADHD-Combined, and ADHD-Inattentive groups. The figure displays aperiodic-adjusted power during EO condition, computed using a canonical frequency range (theta: 4-8 Hz; beta: 13-30 Hz) and TBR. Electrodes labels highlighted on the topographies correspond to the six regions of interest derived from literature used across different branches of multiverse analysis. The power spectra and aperiodic signal were computed by averaging across all electrodes within each respective region of interest. Note. HC = healthy controls. Comb = Combined. In = Inattentive.

##### 1.2.2.2.2. 1/f-uncorrected power

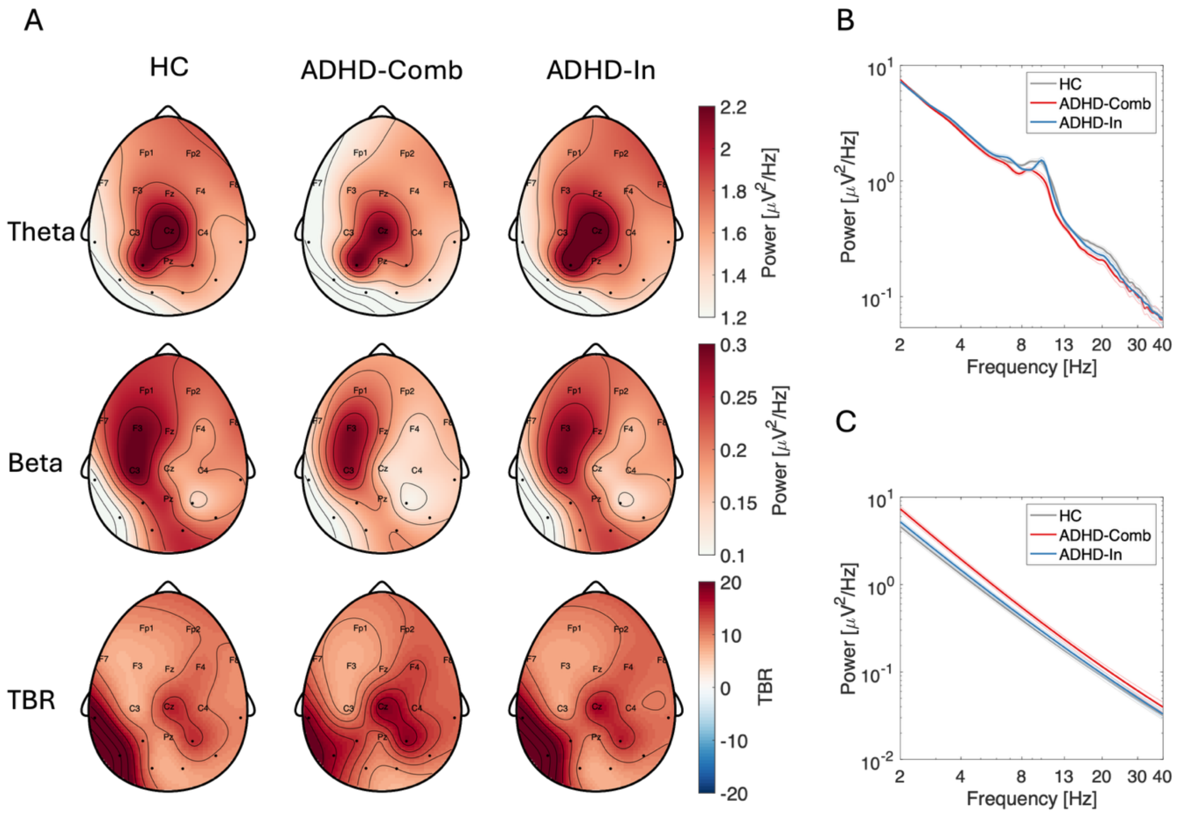


Supplementary Figure 8. Neurophysiological data from the validation sample. (A) Scalp topographies, (B) 1/f-uncorrected power spectra and (C) aperiodic signal for HC, ADHD-Combined, and ADHD-Inattentive groups. The figure displays 1/f-uncorrected power during EO condition, computed using a fixed frequency range (theta: 4-8 Hz; beta: 13-30 Hz) and TBR. Electrodes labels highlighted on the topographies correspond to the six regions of interest derived from literature used across different branches of multiverse analysis. The power spectra and aperiodic signal were computed by averaging across all electrodes within each respective region of interest. Note. HC = healthy controls. Comb = Combined. In = Inattentive.

#### 1.2.2.3. Multiverse analysis results

To evaluate the robustness of effects in the validation sample, we conducted a multiverse analysis using the same regression model:

$$TBR \sim Group * Age * Gender * IAF$$

Group was coded identically as in the HBN sample, with HC as the reference category, and ADHD-Inattentive and ADHD-Combined as the comparisons groups. Gender was coded with males as the reference group. Supplementary Table 10 summarizes the number of significant effects for each term across the 144 model specifications. The multiverse analysis in the validation sample included 144 rather than 576 specifications, as information on comorbid diagnoses and medication status was not available (144 * 2 * 2 = 576 in the HBN sample).

Supplementary Table 10. Results of the multiverse analysis in the validation sample showing the proportion of significant effects across 144 universes.

| | ***Significantly Positive*** | ***Significantly Negative*** | ***Not Significant*** | ***Total*** |
|---|---|---|---|---|
| ADHD[IN] | 2 (1.39 %) | 11 (7.64 %) | 131 (91.00 %) | 144 |
| ADHD[IN] * Age | 1 (0.69 %) | 16 (11.10 %) | 127 (88.20 %) | 144 |
| ADHD[IN] * Gender[Female] | 4 (2.78 %) | 1 (0.69 %) | 139 (96.50 %) | 144 |
| ADHD[IN] * IAF | 0 (0.00 %) | 10 (6.94 %) | 134 (94.10 %) | 144 |
| ADHD[IN] * Age * Gender[Female] | 1 (0.69 %) | 0 (0.00 %) | 143 (99.30 %) | 144 |
| ADHD[IN] * Age * IAF | 30 (20.80 %) | 0 (0.00 %) | 114 (79.20 %) | 144 |
| ADHD[IN] * Gender[Female] * IAF | 7 (4.86 %) | 0 (0.00 %) | 137 (95.10 %) | 144 |
| ADHD[IN] * Age * Gender[Female] * IAF | 0 (0.87 %) | 5 (3.47 %) | 139 (96.50 %) | 144 |
| ADHD[Comb] | 1 (0.69 %) | 2 (1.39 %) | 141 (97.90 %) | 144 |
| ADHD[Comb] * Age | 4 (2.78 %) | 19 (13.20 %) | 121 (84.00 %) | 144 |
| ADHD[Comb] * Gender[Female] | 0 (0.00 %) | 0 (0.00 %) | 144 (100.00 %) | 144 |
| ADHD[Comb] * IAF | 0 (0.00 %) | 1 (0.69 %) | 143 (99.30 %) | 144 |
| ADHD[Comb] * Age * Gender[Female] | 0 (0.00 %) | 8 (5.56 %) | 136 (94.40 %) | 144 |
| ADHD[Comb] * Age * IAF | 14 (9.72 %) | 6 (4.17 %) | 124 (86.10 %) | 144 |
| ADHD[Comb] * Gender[Female] * IAF | 2 (1.39 %) | 0 (0.00 %) | 142 (98.60 %) | 144 |
| ADHD[Comb] * Age * Gender[Female] * IAF | 0 (0.00 %) | 27 (18.80 %) | 117 (81.20 %) | 144 |

Note. Positive universe = higher TBR in ADHD compared to HC. Negative universe = lower TBR in ADHD compared to HC. IN = Inattentive. Comb = Combined. IAF = Individual alpha frequency.

###### 1.2.2.4. Healthy Controls vs. ADHD-Inattentive Comparison

The results largely mirrored the HBN sample (Supplementary Figure 9). No consistent main effect of ADHD-Inattentive was observed, with only 2 positive and 11 negative universes out of 144 possibilities. Out of those 11 negative universes, 8 were observed when IAF-related frequency band and aperiodic signal. Next, the ADHD-Inattentive * Age interaction resulted in 16 significantly negative universes, which were again observed when using IAF-related frequency band and aperiodic signal (8 out of 24 possible universes; Supplementary Figure 10 A) and 1/f-uncorrected power (8 out of 24 possible universes). Finally, the ADHD-Inattentive * Age * IAF interaction was significantly positive in 30 universes, especially when frequency bands were defined relative to IAF and when using aperiodic signal (15 out of 24 possible universes; Supplementary Figure 10 B) and 1/f-uncorrected power (15 out of 24 possible universes; Supplementary Figure 10 C).

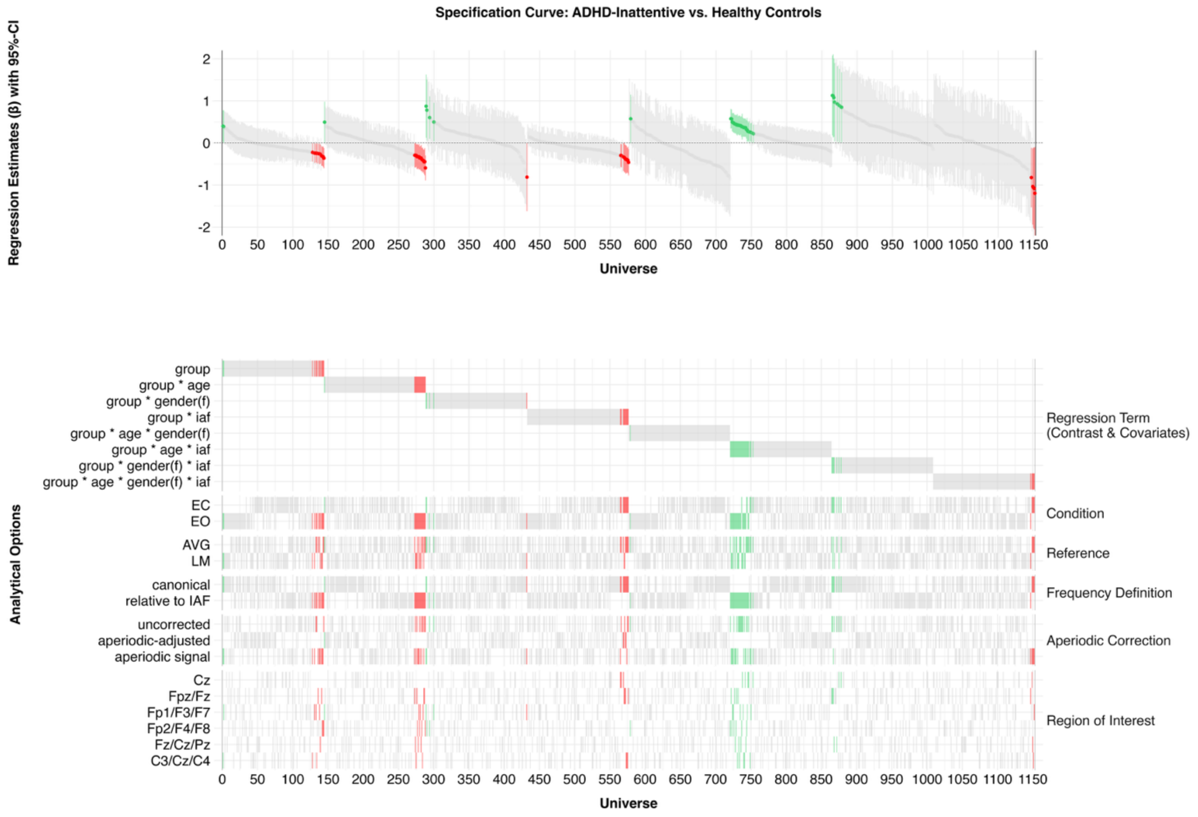


Supplementary Figure 9. Specification curve representing all universes for the healthy control vs. ADHD-Inattentive comparison in the validation sample. The top panel shows regression estimates sorted by effect size with 95% confidence intervals. Statistically significant positive estimates are shown in green, negative in red, and non-significant in gray. The bottom panel maps the analytical choices that produced each estimate. Note. CI = confidence interval. f = female. EC = eyes closed. EO = eyes open. AVG = average reference. LM = linked mastoid reference. canonical = canonical frequency range. relative to IAF = bandwidth relative to individual alpha frequency.

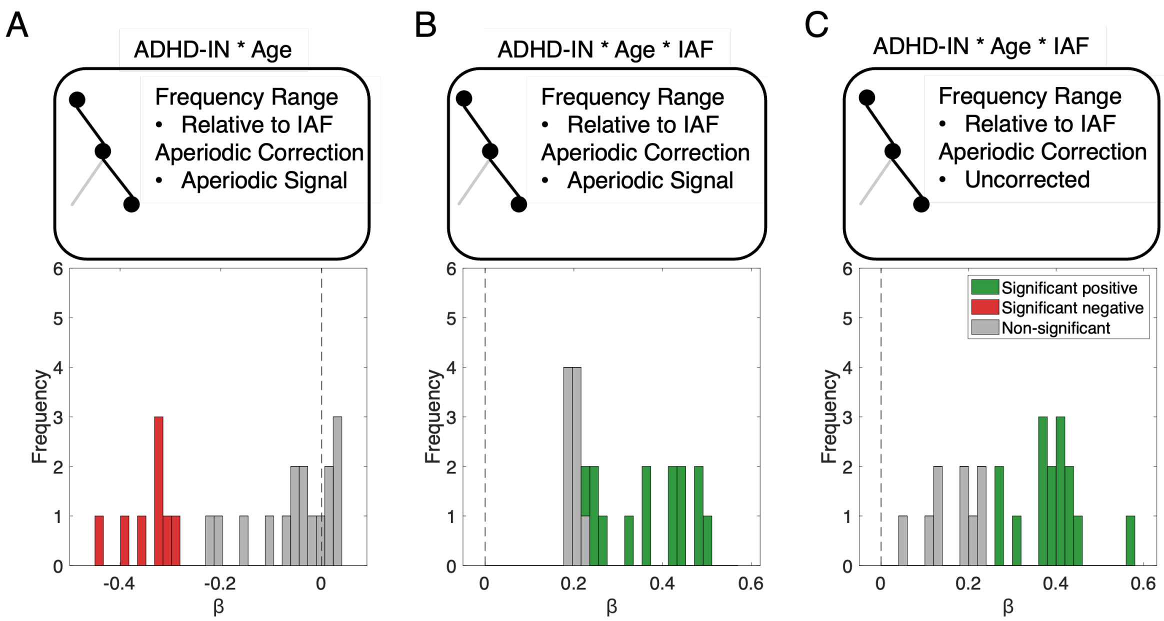


Supplementary Figure 10. Possibility space for the ADHD-Inattentive * Age and ADHD-Inattentive * Age * IAF interactions. (A) Negative ADHD-Inattentive * Age effect: 8 out of 24 significant universes with IAF-relative bands and aperiodic signal. (B) Positive ADHD-Inattentive * Age * IAF effect: 15/24 significant under aperiodic signal with IAF-relative bands (C) Positive ADHD-Inattentive * Age * IAF effect: 15/24 significant with IAF-relative bands and uncorrected power. Note. The significant positive estimates are highlighted green, significant negative in red and non-significant in grey.

##### 1.2.2.5. Healthy Controls vs. ADHD-Combined Comparison

Lastly, we computed the contrast between the HC and ADHD-Combined groups. The specification curve for this analysis is shown in Supplementary Figure 11. In the HC vs. ADHD-Combined contrast, we observed an ADHD-Combined * Age interaction that was significantly negative in 19 universes, specifically in 12 out of 24 specifications involving IAF-related bands and the 1/f-uncorrected signal (Supplementary Figure 12 A). Furthermore, the ADHD-Combined * Age * IAF interaction was significantly positive in 14 universes, all of which were based on IAF-related band definitions and the 1/f-uncorrected signal (Supplementary Figure 12 B). Finally, the ADHD-Combined * Age * Gender * IAF interaction was significantly negative in 27 out of 144 universes, with 13 of these effects arising from specifications using IAF-related bands and the 1/f-uncorrected signal (Supplementary Figure 12 C).

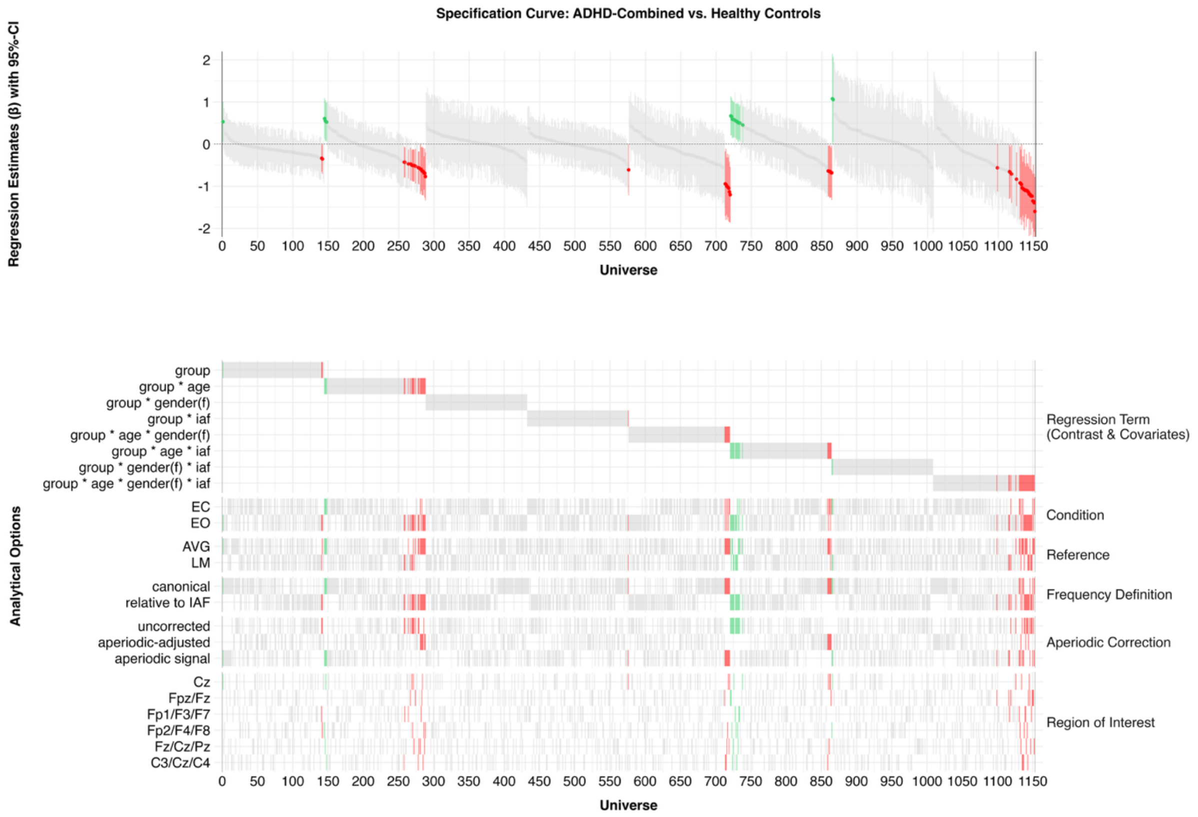


Supplementary Figure 11. Specification curve representing all universes for the healthy control vs. ADHD-Combined comparison in the validation sample. The top panel shows regression estimates sorted by effect size with 95% confidence intervals. Statistically significant positive estimates are shown in green, negative in red, and non-significant in gray. The bottom panel maps the analytical choices that produced each estimate. Note. CI = confidence interval. f = female. EC = eyes closed. EO = eyes open. AVG = average reference. LM = linked mastoid reference. canonical = canonical frequency range. relative to IAF = bandwidth relative to individual alpha frequency.

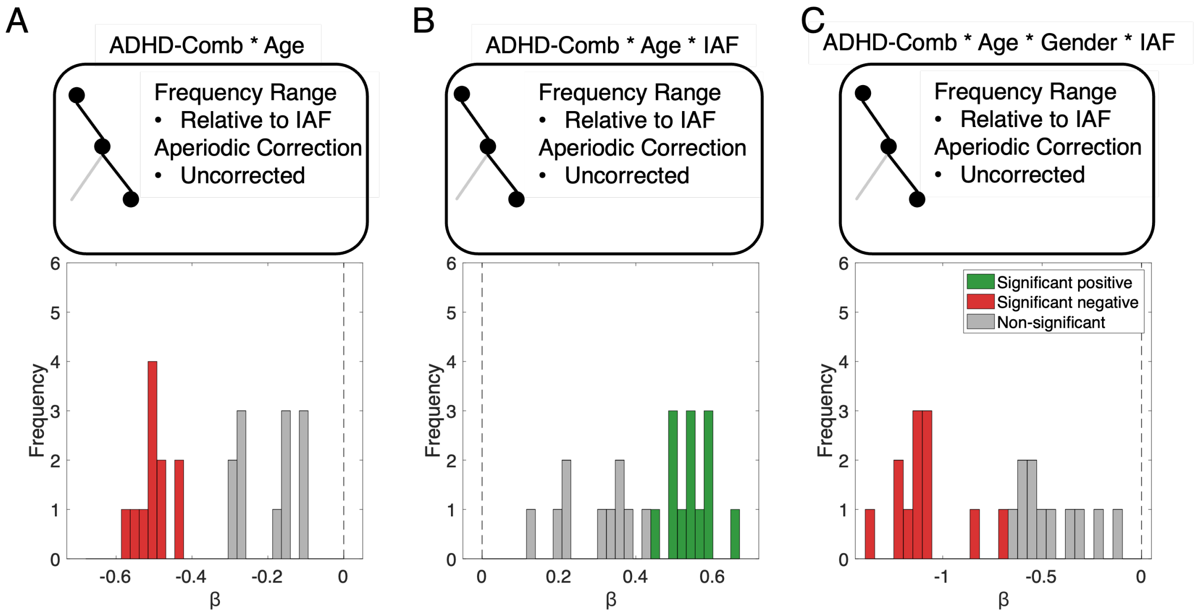


Supplementary Figure 12. Possibility space for the ADHD-Combined * Age, ADHD-Combined * Age * IAF and ADHD-Combined * Age * Gender * IAF interactions. (A) Negative ADHD-Combined * Age effect: 12 out of 24 significant universes with IAF-relative bands and uncorrected power. (B) Positive ADHD-Combined * Age * IAF effect: 14/24 significant under aperiodic signal with IAF-relative bands (C) Positive ADHD-Combined * Age *Gender * IAF effect: 13/24 significant with IAF-relative bands and uncorrected power. Note. The significant positive estimates are highlighted green, significant negative in red and non-significant in grey.

##### 1.2.2.6. Proportion Plots Across All Analytical Specifications

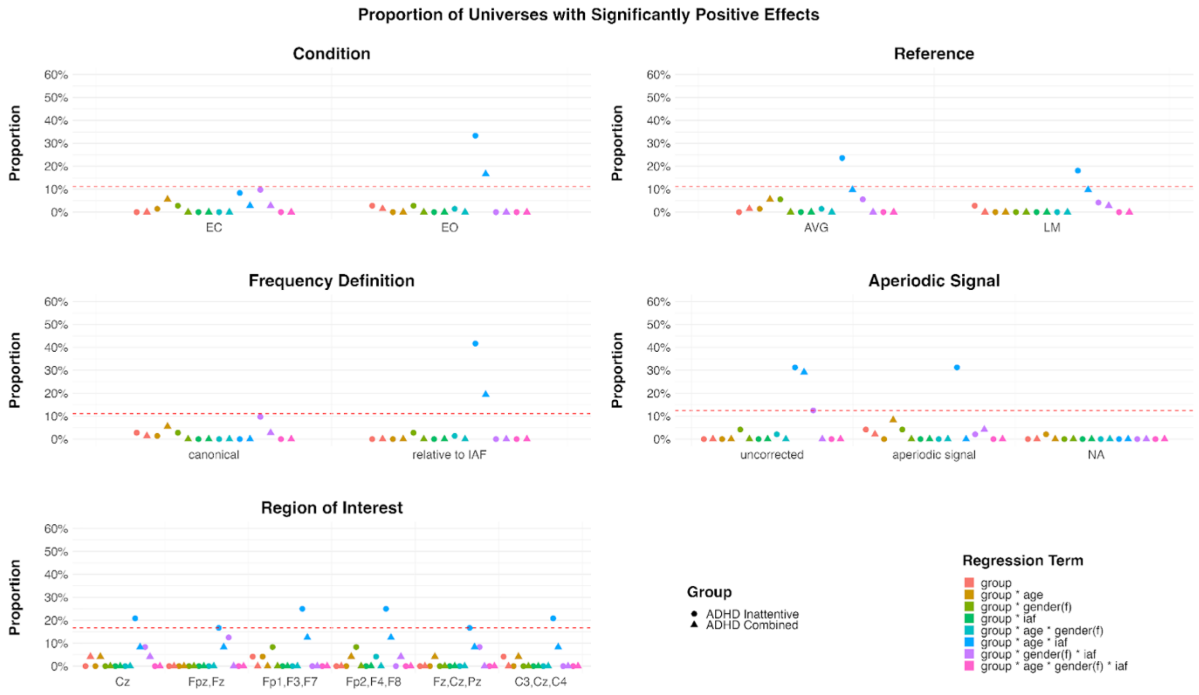


Supplementary Figure 13. Proportion of specifications showing a significantly positive effect by regression term and ADHD subtype across all analytical specifications. Each level of Condition, Reference, Frequency Definition includes 72 analytical universes. The Aperiodic Signal factor includes 48 universes per level, and the Region of Interest includes 24 universes per level. The red line in the plot indicates the exact proportion of significant results required for the binomial test to reach statistical significance ($p < .05$) for each subset size.

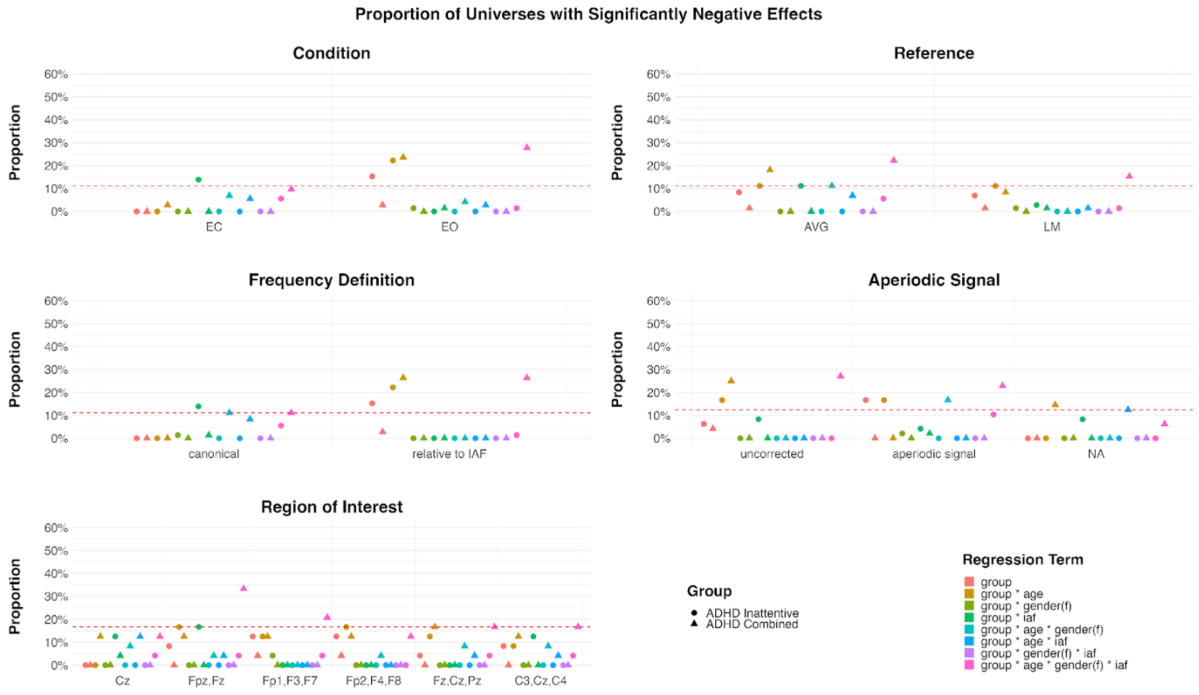

Supplementary Figure 14. Proportion of specifications showing a significantly negative effect by regression term and ADHD subtype across all analytical specifications. Each level of Condition, Reference, Frequency Definition includes 72 analytical universes. The Aperiodic Signal factor includes 48 universes per level, and the Region of Interest includes 24 universes per level. The red line in the plot indicates the exact proportion of significant results required for the binomial test to reach statistical significance ($p < .05$) for each subset size.
